# Epidemiology and burden of respiratory syncytial virus in Italian adults: A systematic review and meta-analysis

**DOI:** 10.1101/2024.01.11.24301142

**Authors:** Alexander Domnich, Giovanna Elisa Calabrò

## Abstract

**Objective:** Respiratory syncytial virus (RSV) is a common respiratory pathogen not only in children, but also in adults. Country-specific data on the epidemiology and burden of disease are essential for policy decisions. In view of a recent authorization of adult RSV vaccines, we aimed to comprehensively collect and assess evidence on the epidemiology and burden of RSV in Italian adults.

**Methods:** A systematic literature review was conducted according to the available guidelines. Random-effects proportional meta-analysis was performed to obtain pooled estimates and the observed heterogeneity was investigated by using both subgroup and meta-regression analyses.

**Results:** A total of 35 studies were identified. RSV seasonal attack rates ranged from 0.8 ‰ in community-dwelling older adults to 10.9% in hematological outpatients. On average, 4.5% (95% CI: 3.2–5.9%) of respiratory samples tested positive for RSV. This positivity prevalence was higher in older adults (4.4%; 95% CI: 2.8– 6.3%) than in working-age adults (3.5%; 95% CI: 2.5–4.6%) and in outpatient (4.9%; 95% CI: 3.1–7.0%) than inpatient (2.9%; 95% CI: 1.5–4.8%) settings. Study location and sample size were also significant predictors of RSV detection frequency. The pooled estimate of in-hospital mortality was as high as 7.2% (95% CI: 4.7– 10.3%). However, other important indicators of the diseases burden, such as complication and hospitalization rates, are missing.

**Conclusion:** RSV poses a measurable burden on Italian adults, especially those of older age and with some co-morbidities. Policy makers should give priority to health technology assessment of the novel RSV vaccines.

## 1. Introduction

Together with seasonal influenza, respiratory syncytial virus (RSV) is a leading cause of respiratory infections and is responsible for a significant socioeconomic burden in all age groups, especially at the extremes of age [1,2]. A recent modelling study [3] has estimated that on average, a total of 158,229 RSV-associated hospitalizations among European adults occur annually and 92% of these concerns the elderly. Contrary to young children, mortality attributable to RSV increased in both working-age and older adults [4].

Several systematic reviews and meta-analyses (SRMAs) [5–10] have investigated global epidemiology and burden of RSV in (older) adults. These reviews have advocated high incidence, hospitalization, mortality and case-fatality rates of RSV [5–10], which seem similar to seasonal influenza [9], and pointed out a substantial case under-ascertainment [10]. Moreover, there was a large between-country variation in estimates of the burden of disease (BoD) [6], which is driven by numerous factors, from climatic conditions [11] to the features of surveillance systems in place [12]. Notably, the available SRMAs [5–10] have identified only up to six studies conducted in Italy.

Two vaccines have been recently authorized to prevent lower respiratory tract disease (LRTD) caused by RSV in adults aged ≥60 years [13]. This age indication will be likely extended to younger individuals in the upcoming years. Some countries have already established RSV immunization policies. In the United States (US), a single dose of RSV vaccine is recommended to adults ≥60 years, as a part of shared clinical decision-making between patient and healthcare provider [14]. In the United Kingdom (UK), RSV vaccination is currently advised for older adults aged ≥75 years, being the most cost-effective option [15]. By contrast, as of November 2023, no recommendations have been issued in Italy.

Understanding country-specific BoD is a key driver for policy decisions on the introduction of new vaccines [16]. A systematic appraisal of the burden of RSV enables policy makers, health professionals and other relevant stakeholders to make informed decisions regarding the recently available vaccines. In this regard, SRMAs on different BoD indicators are important in the description of spatiotemporal distribution and variations between population subgroups potentially targeted by the novel preventive measures [17,18]. Systematically appraised country-specific BoD indicators are also essential for all types of pharmacoeconomic models.

In Italy, a recent review [19] has assessed RSV BoD in pediatric outpatients, also with the aim to inform decision makers on the recent availability of a monoclonal antibody for RSV prevention in neonates. By including six studies, the authors found that 18–41% of children with respiratory infections were positive for RSV. Conversely, no reviews have systematically assessed burden of RSV in Italian adults. Indeed, RSV epidemiology is highly age-dependent, which hinders transferability of pediatric estimates to older populations. Furthermore, as we mentioned earlier, the available global-level reviews [5–10] were able to identify only a limited number of Italian studies. In this SRMA, we aimed to comprehensively collect and analyze available data on RSV epidemiology and BoD in Italian adults with the ultimate goal of informing and supporting National and local decision makers on the planification and implementation of vaccination strategies.

## 2. Materials and methods

### 2.1. Reporting standards and protocol

PRISMA (preferred reporting items for SRMAs) statement [20] was adopted as a reporting standard (S1 Table). Methodological guidance for systematic reviews of observational epidemiological studies reporting prevalence and cumulative incidence data developed by the Joanna Briggs Institute (JBI) [21] was also consulted. The study protocol was prospectively registered [22] and no amendments to the original protocol were made.

### 2.2. Eligibility criteria

All types of observational studies and published in any modality (e.g., peer-reviewed article, preprint, conference abstract, etc.) were potentially eligible. The CoCoPop (condition, context, and population) approach [21] was used to formulate the inclusion criteria. In particular, the condition of interest was RSV infection detected by any laboratory technique, including reverse-transcription polymerase chain reaction (RT-PCR), culture, immunofluorescence assay (IFA) and rapid antigen tests. Moreover, RSV-specific International Statistical Classification of Diseases and Related Health Problems (ICD) diagnosis codes (see below) were also considered a good proxy for the true RSV infection, since their specificity is as high as 99.6–99.8% [23,24]. For the context, we considered studies conducted in Italy, in any setting (outpatient, inpatient or mixed), time and calendar period. Population consisted of adults defined as individuals aged ≥14 years, independently of their health conditions. Any reason or clinical entity [e.g., influenza-like illness (ILI), acute respiratory infection (ARI), severe ARI (SARI), clinical request for differential diagnosis] that triggers collection of biological samples (upper respiratory tract specimens including naso/oropharyngeal and nasal swabs; lower respiratory tract specimens including sputum, bronchoaspirates and bronchoalveolar lavage fluids) was eligible.

The following were set as exclusion criteria: (i) modeling, pharmacoeconomic and similar studies with no original data; (ii) insufficient data on RSV; (iii) studies on general population with no separate data for adults; (iv) multi-country studies with no separate data for Italy; (v) redundant publications.

### 2.3. Study endpoints

RSV attack rate (cumulative incidence) was defined as the occurrence of laboratory-confirmed RSV detection in a population (symptomatic, asymptomatic or both) and in a specific period. RSV positivity prevalence was defined as proportion of RSV detections to the number of subjects tested. Prevalence of viral co-detections was described as number of samples tested positive for both RSV and any other respiratory virus to the total number of RSV-positive samples. Case-complication rate was defined as proportion of subjects who tested positive for RSV and developed ≥1 complication, such as pneumonia, exacerbation of chronic obstructive pulmonary disease (COPD), asthma, congestive heart failure, and other. As for drug use indicators, we considered frequency of antibiotic prescriptions among RSV-positive subjects. For what concerns inpatient outcomes, crude hospitalization, case-hospitalization (i.e., proportion of RSV-positive individuals who were hospitalized) rates, length of stay [mean with standard deviation (SD) or median with interquartile range (IQR)] and frequency of admission to intensive care units (ICUs) were of interest. Analogously, crude, in-hospital, 30-day mortality and case-fatality rates were eligible. For the indicators based on hospital discharge records (HDRs) or death certificates, we considered only RSV-specific codes, namely RSV pneumonia (ICD-9: 480.1; ICD-10: J12.1), acute bronchiolitis due to RSV (ICD-9: 466.11; ICD-10: J21.0), acute bronchitis due to RSV (ICD-10: J20.5), and RSV as the cause of diseases classified elsewhere (ICD-9: 079.6; ICD-10: B97.4).

When possible, all study endpoints were described overall, by age-group (working-age and older adults), RSV subtype (A and B) and season.

### 2.4. Search strategy

The automatic search was performed on 22 November 2023 in the following databases: (i) MEDLINE via Ovid; (ii) Biological Abstracts via Ovid; (iii) Global Health via Ovid; (iv) Scopus and (v) Web of Science. In order to increase sensitivity, no filters or other restrictions (e.g., language or publication year) were applied. The search script considered both MeSH (medical subject headings) and text-wide terms and is reported in S2 Table.

We then performed a manual search through several modalities. First, the reference lists of the available global-level SRMAs [5–10] were checked. Second, a backward cross-reference checking of the included studies was carried out. Third, a forward citation search by using Google Scholar (https://scholar.google.com/) was conducted, as this search engine is better suited for identifying grey literature sources [25]. Fourth, periodic reports of the Italian surveillance reports on influenza and other respiratory viruses (https://respivirnet.iss.it) were examined. Finally, we screened abstract books and proceedings of some relevant conferences, including ECCMID (European Society of Clinical Microbiology and Infectious Diseases), ESWI (European Scientific Working Group on Influenza) and ReSViNET.

### 2.5. Study selection

Results produced by the automatic search were merged into a single spreadsheet and duplicates were removed. The resulting list of unique records underwent screening by assessing titles and/or abstracts and clearly irrelevant citations were discarded. Full texts of potentially pertinent publications were then downloaded and assessed for the above-described eligibility criteria. Study selection was finalized by performing the manual search, as described earlier. The entire process of study selection was performed by both reviewers, each working independently; eventual disagreements were solved by consensus.

### 2.6. Data extraction and abstraction

The following data were extracted: (i) full citation record; (ii) study location; (iii) study period; (iv) study design; (v) study setting; (vi) main characteristics of the study population; (vii) sample size; (viii) funding source; (ix) eligibility criteria and case definitions; (x) methods used for case ascertainment; (xi) numerators and denominators used to compute the endpoints of interest described above; (xii) other potentially relevant information.

On the basis of period, studies were dichotomized on whether they overlapped with the COVID-19 pandemic, which had a significant impact on the circulation of RSV and other respiratory viruses. In particular, the northern hemisphere winter season 2020/2021 was characterized by a very limited circulation of RSV [19,26]. Estimates for that season were extracted, but not included in the quantitative synthesis. Starting from the 2021/2022 season, RSV returned to the epidemiological scene [26] and therefore estimates from 2021/2022 onwards were fully considered. Multi-season studies that reported separate seasonal data were considered as distinct estimates [27]. Moreover, we distinguished between year-around studies and those conducted during a typical RSV season, in which the probability of RSV detection is much higher [28]. We defined RSV epidemic season as a period between October and April [26].

On the basis of sample size, studies were median split. Regions of were categorized into three macro-areas of North (Aosta Valley, Liguria, Lombardy, Piedmont, Emilia-Romagna, Friuli-Venezia Giulia, Trentino-South Tyrol, Veneto), Center (Lazio, Marche, Tuscany, Umbria) and South (Abruzzo, Apulia, Basilicata, Calabria, Campania, Molise, Sicily, Sardinia).

Missing data on relevant numerators and/or denominators were handled as follows. First, the corresponding author was contacted for clarification. In case of no reply, these data were imputed from the available percentages and/or by extracting data from figures using the WebPlotDigitizer v.4.6 software (https://automeris.io/WebPlotDigitizer). Data were extracted by AD and then validated by GEC.

### 2.7. Critical appraisal

The JBI checklist for prevalence studies [21] was used to assess quality of the included studies. It was assessed independently by both reviewers and eventual conflicts were solved by consensus. Item 9 of the JBI checklist on the response rate was judged irrelevant for this study. Owing to a limited information available, risk of bias of conference abstracts, letters to the editor, short communications and similar was not assessed.

### 2.8. Data synthesis

Tabulated data were first reassumed qualitatively and by visualizing forest plots. For quantitative synthesis, a proportional meta-analysis was undertaken according to the available recommendations [18,21]. As heterogeneity was expected to be high, random-effects (RE) models with double arcsine transformation to stabilize variances were used. Pooled estimates were expressed as proportions with 95% Clopper-Pearson confidence intervals (CIs). Heterogeneity was quantified by means of the τ^2^ and I^2^ statistics. Notably, high I^2^ values do not necessarily mean that the data are inconsistent, as true heterogeneity is expected in prevalence estimates due to spatiotemporal differences [18]. The 95% prediction intervals (PIs) were also computed. As recommended, publication bias was not formally assessed, since there is no consensus about what a positive result in meta-analyses of proportions is [18].

To investigate the sources of heterogeneity across studies, both subgroup and meta-regression analyses were performed. In particular, the subgroup analysis was performed by age-group (working-age and older adults), setting (outpatient, inpatient and mixed) and study period in relation to the COVID-19 pandemic (before and after the 2020/2021 season). The meta-regression modeling was then conducted to examine the influence of study characteristics on the RSV-related endpoints. This latter was performed only when ≥10 estimates were available [29]. A leave-one-out sensitivity analysis was finally conducted to check the robustness of pooled estimates.

Meta-analysis was performed in R (R Foundation for Statistical Computing; Vienna, Austria) package “Meta” v. 6.5-0.

## 3. Results

### 3.1. Characteristics of the included studies

The automatic search generated 312 records, of which 171 were duplicates. Following screening of 141 records, 37 were judged potentially eligible. Twelve studies were excluded with reasons and are reported in S3 Table. Manual search identified further 12 studies and therefore the final list was composed of 35 studies corresponding to 37 publications [30–66]. Notably, results relative to a retrospective cohort (henceforth referred to as “Boattini 2021–2023”) were presented in three different publications [64–66]. Following correspondence with the corresponding author, who provided additional data, it was decided to include all these records. Moreover, relevant data (not reported within the article) were also obtained from other two corresponding authors [55,59]. The entire study selection process is reported in Fig 1.

**Fig 1.**
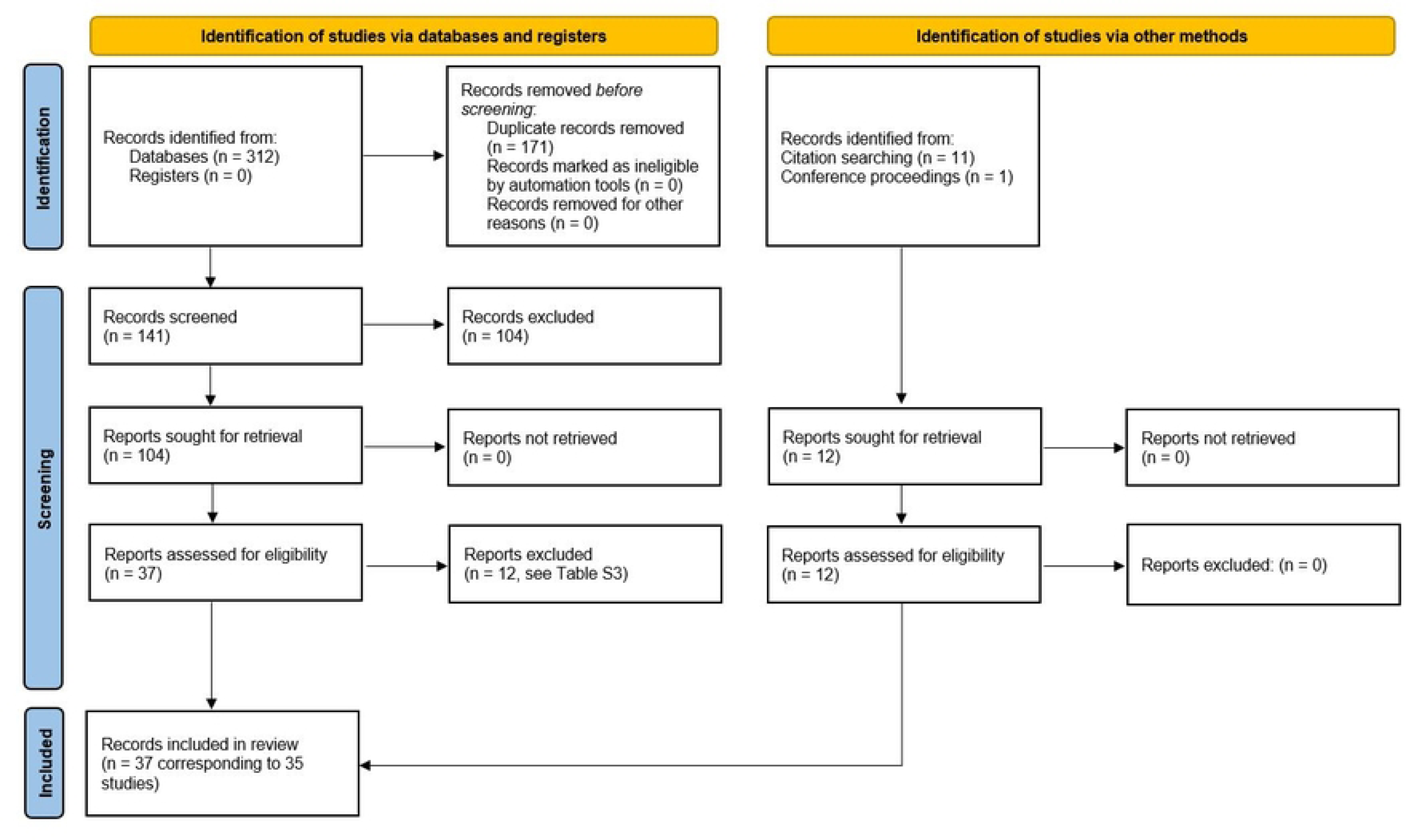
PRISMA flow diagram of the study selection process.

Most (83%; 29/35) studies included were full-length articles, while the remaining six were conference abstracts [45,47], letters to editor [43,57] and short papers [31,39]. Principal characteristics of the studies are summarized in Table 1. Briefly, the studies covered a period from the 2001/2002 season to the 2022/2023 season and most (63%; 22/35) were conducted in the north of Italy. Approximately half (49%; 17/35) of the studies investigated more than one RSV season and the period of nine (26%) studies overlapped with the COVID-19 pandemic. Design of the majority of studies was judged cross-sectional (54%; 19/35) or surveillance (29%; 10/35) and the median sample size was 328 (range 43–28,500) patients. The study population was composed of outpatients, inpatients and mixed groups in 20% (7/35), 37% (13/35) and 31% (11/35) of studies respectively; the remaining four (11%) studies were focused on hematological [43–45] and cystic fibrosis [57] in-/outpatients. The setting of these latter four studies was therefore categorized as “immunocompromised patients”. All but one studies used at least one laboratory assay (mostly RT-PCR) for RSV detection. The remaining study [35] analyzed RSV-specific HDRs. Within laboratory-based studies (n = 34), there was a high level of heterogeneity in terms of clinical entity triggering the specimen collection, especially in mixed and hospital settings. Six out 7 (86%) outpatient studies enrolled ILI patients. ILI and SARI were more frequently defined according to the European criteria (S4 Table). Out-of-season samples were frequent (38%; 13/34) and most studies analyzed only (56%; 19/34) upper respiratory tract specimens. The majority (74%; 26/35) of studies were funded by public institutions.

**Table 1.**
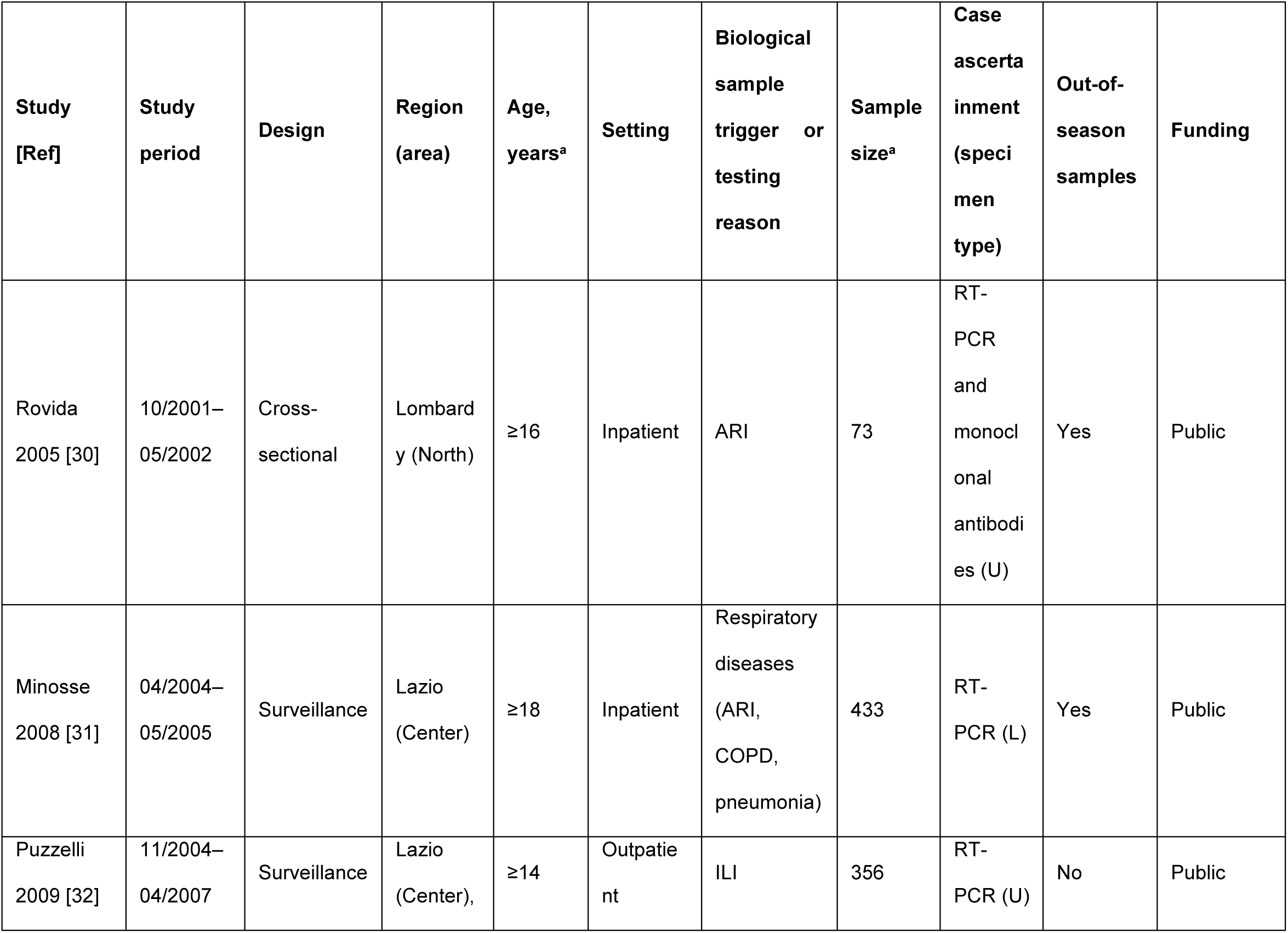
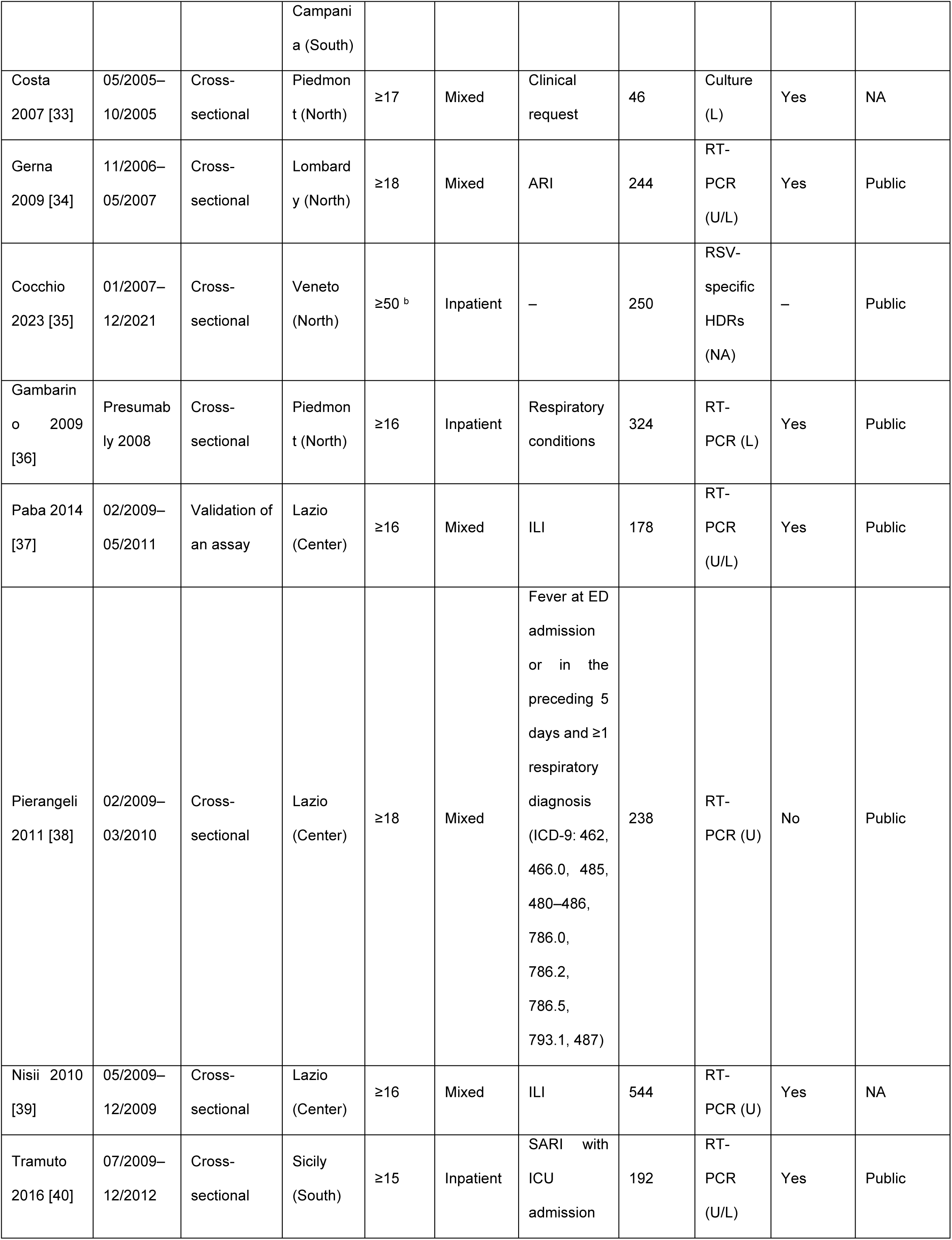
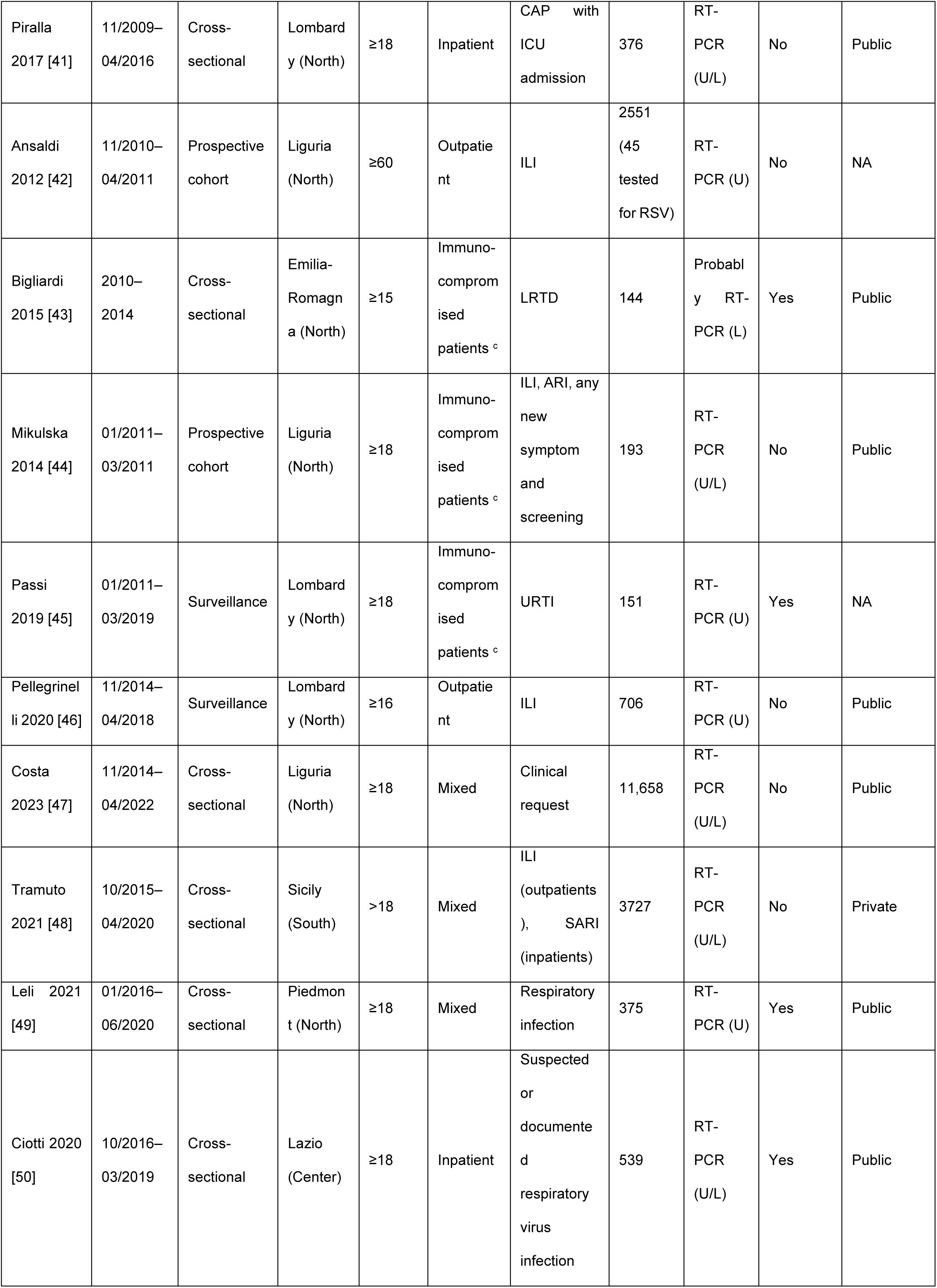
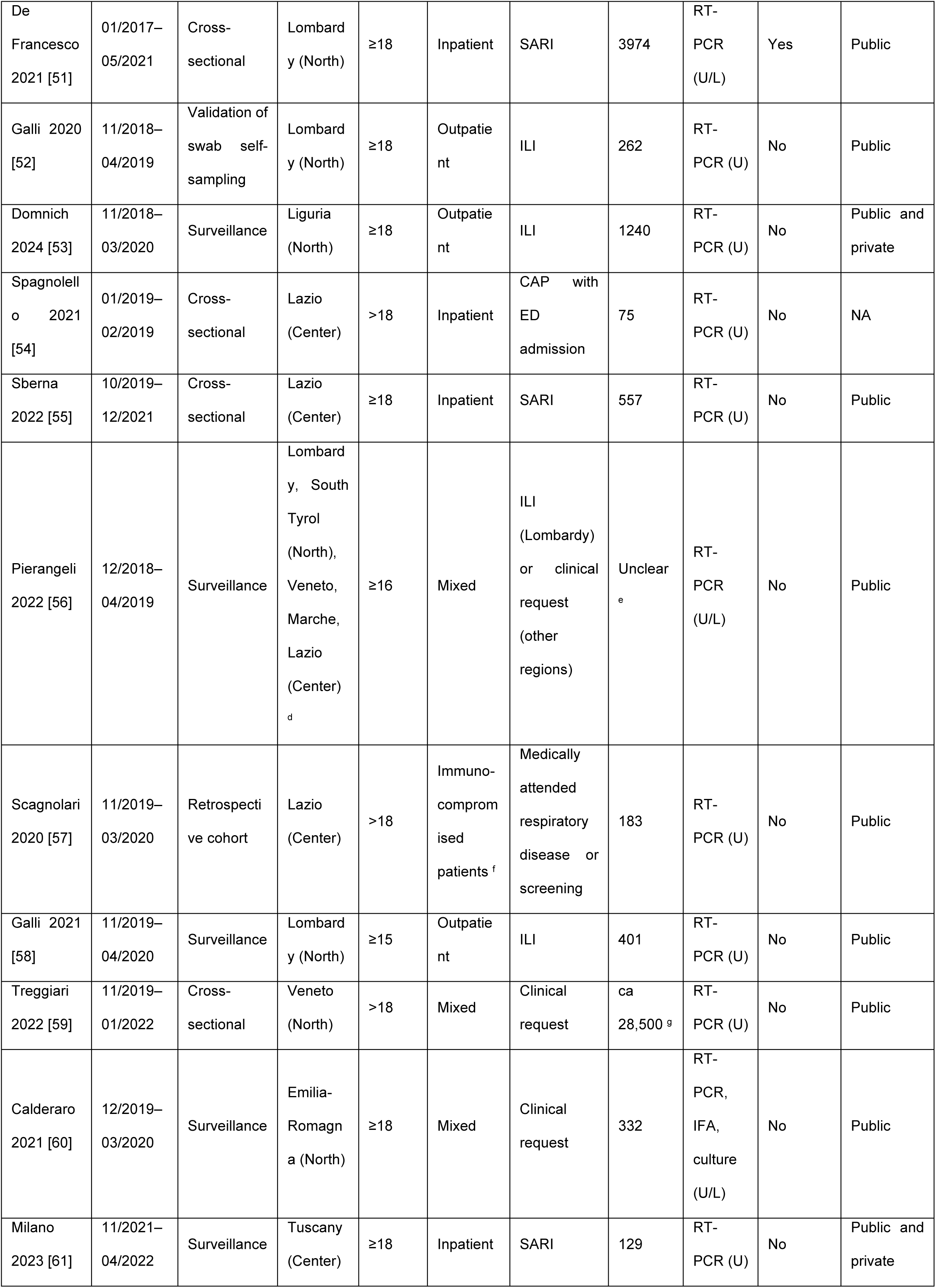
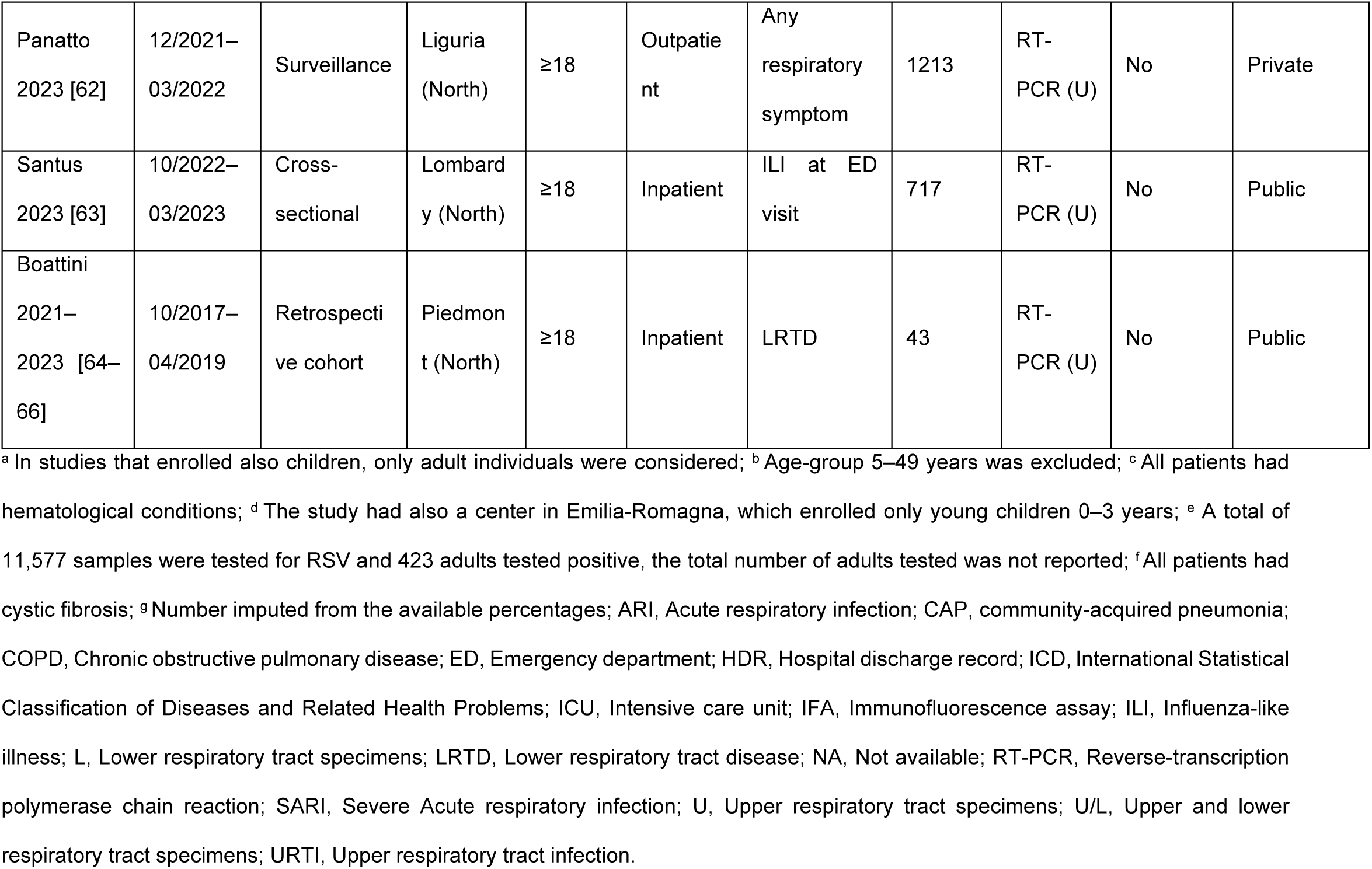
Characteristics of the studies included.

Fourteen [32,38,40,41,44,46,48,51,53,58,61–66] out of 29 full-length articles were judged at low risk of bias, while the remaining 15 studies [30,33–37,42,49,50,52,54–56,59,60,64–66] had at least one possible source of bias (S5 Table). The most frequent criticalities concerned insufficient description of patients and setting, inappropriate sample frame and small sample size.

### 3.2. RSV attack rate

Three cohort studies [42,44,57] allowed to determine RSV attack rates. The first one [42] consisted of a cohort of 2551 community-dwelling adults aged ≥60 years who were actively surveilled for ILI between November 2010 and April 2011. A total of 45 ILI cases were prospectively identified and these cases were tested in RT-PCR. Other 63 ILI cases were identified retrospectively and thus not tested in RT-PCR. Two out of 45 samples tested positive for RSV, giving a symptomatic attack rate of 0.8 ‰ (95% CI: 0.1–2.8‰). If corrected for under-testing, the cumulative incidence would rise to 1.9 ‰ [42]. Mikulska et al. [44] followed for three months (from January to March 2011) 193 hematological outpatients. A total of 21 swabs tested positive for RSV, with an overall attack rate of 10.9% (95% CI: 6.9–16.2%). Of RSV-positive subjects, 18 (85.7%) were symptomatic, while the remaining three (14.3%) patients were asymptomatic [44]. In a retrospective cohort of adult (>18 years) subjects affected by cystic fibrosis [57] eight out of 183 patients tested positive for RSV between November 2019 and March 2020, giving an attack rate of 4.4% (95% CI: 1.9–8.4%). Owing to different study populations, a pooled analysis of these three studies was judged unfeasible.

### 3.3. Prevalence of RSV positivity

A total of 85 RSV positivity prevalence estimates were extracted from 32 studies, of which 42 (S6 Table), 21 (S7 Table) and 22 (S8 Table) concerned adults of any age, working-age and older adults, respectively. Seven estimates from three studies [51,55,59] covered exclusively the 2020/2021 season and were excluded from the pooled analysis.

As shown in Fig 2, the positivity rate in adults of any age ranged from 0.2% to 35.7%. The RE pooled estimate was 4.5% (95% CI: 3.2–5.9%). As expected, the heterogeneity was high (I^2^ = 93.2%) and the 95% PI was 0.0– 15.9%. Immunocompromised patients showed the highest RSV positivity prevalence of 11.5%. Compared with outpatients (4.9%), inpatients had lower prevalence of RSV (2.9%). Studies conducted in both settings showed an intermediate pooled estimate (3.7%). Omission of single studies did not alter significantly the observed pooled proportions.

**Fig 2.**
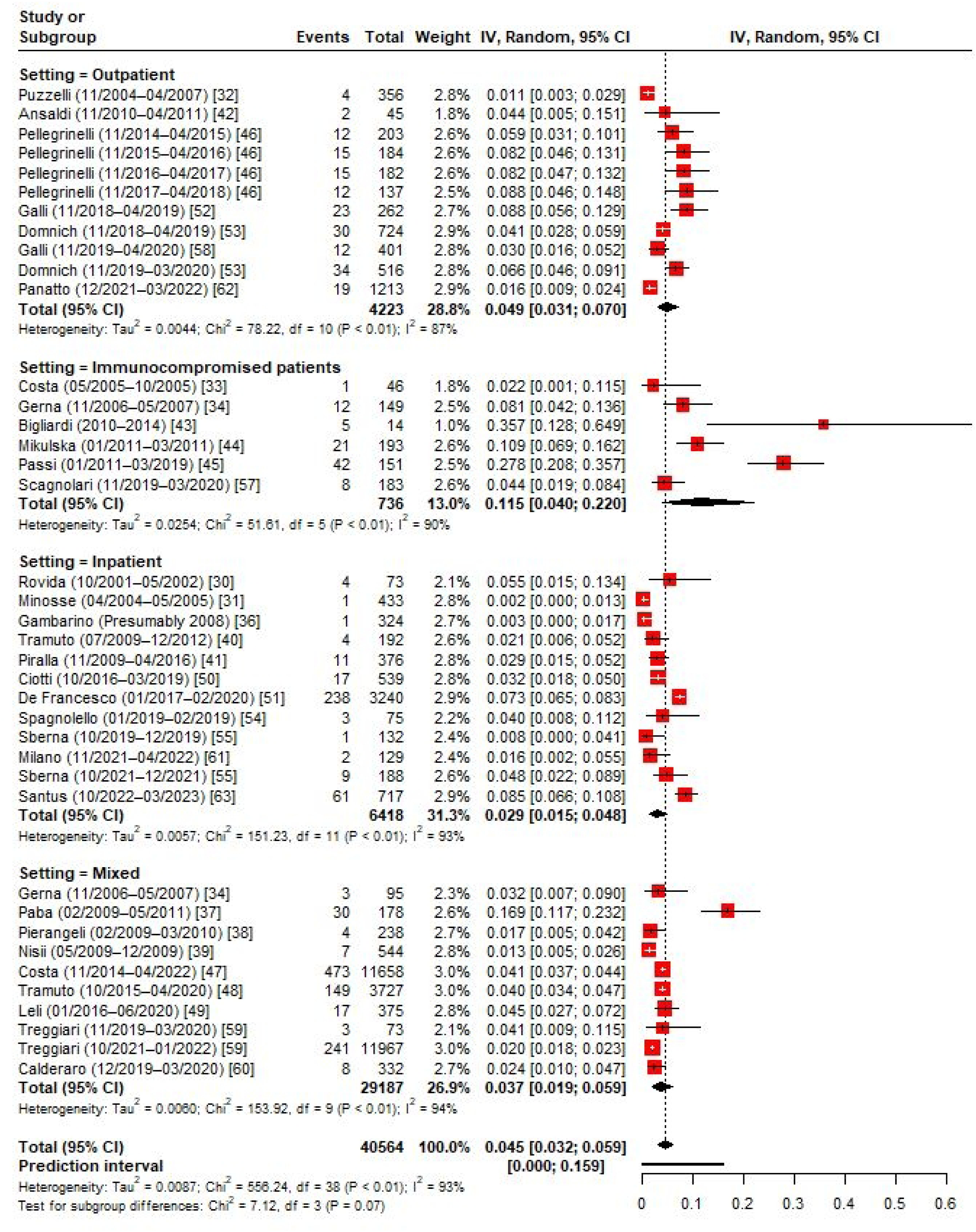
**Forest plot of the random-effects model on RSV positivity prevalence among Italian adults of any age, by setting.**

In the subgroup analysis by age-group (Fig 3), working-age adults showed lower (3.5%) RSV prevalence than older adults (4.4%). RSV detection was more frequent in outpatients than inpatients in both age-specific RE models. RSV detection was also higher in studies conducted before the COVID-19 pandemic than among those performed during the pandemic (i.e., seasons 2021/2022 and 2022/2023) (S1 Fig).

**Fig 3.**
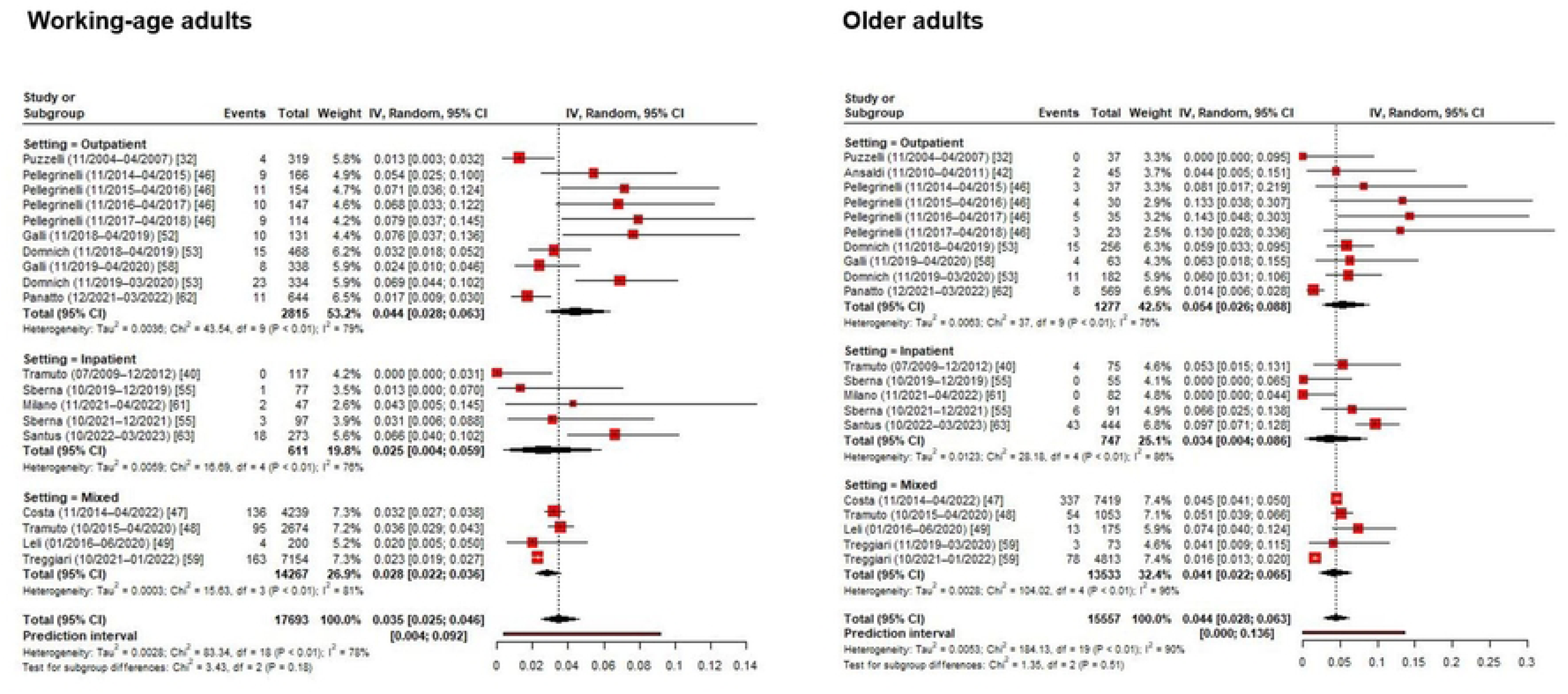
**Forest plot of the random-effects model on RSV positivity prevalence among Italian working-age and older adults, by setting.**

A meta-regression analysis was conducted to investigate sources of the heterogeneity observed (S9 Table). Apart from the study setting, geographic area and sample size explained some variance in the reported proportions. In particular, compared with northern regions (5.5%), RSV prevalence was as twice as lower in central Italy (2.7%) (S2 Fig). Studies that enrolled <300 patients reported significantly higher prevalence (6.3%) estimates than larger studies (3.0%) (S3 Fig). Publication year, public funding, number of seasons, inclusion of out-of-season samples, specimen type and risk of bias were not associated with the reported RSV detection rate (S9 Table).

Data on RSV subtypes was reported in nine studies [39,40,44,46–48,53,56,62]. In these studies, both subtypes co-circulated, although a relative dominance on one subtype over another varied by study period (S10 Table). On average, from 2009 to 2022 both subtypes were detected in almost equal proportions (RE model estimate for RSV B: 56.3%; 95% CI: 44.1–68.1%; I^2^ = 88.4%) (S4 Fig).

Sanger sequencing was performed in three studies [46,56,62] conducted between 2014/2015 and 2021/2022 seasons. During the 2014/15 and 2015/16 seasons, both NA1 and ON1 genotypes of RSV A were circulating [46]. Conversely, starting from the 2017/2018 season all RSV A strains clustered within the ON1 genotype [46,56,62]. All RSV B strains belonged to the BA genotypes [46,56,62].

### 3.4. Prevalence of viral co-detections

Thirteen studies reported data on viral co-detections [30,31,36,37,40,47,51,53,54,60–63]. However, only five estimates [37,47,51,53,63] were based on a sufficient (≥30) number of RSV-positive samples and were therefore less prone to the small study effect (S11 Table). In these studies, 2.9–18.6% of RSV-positive specimens tested also positive for other respiratory viruses. In the RE model (S5 Fig), the pooled estimate was 8.6% (95% CI: 3.2–15.9%) and the heterogeneity was high (I^2^ = 92.0%). Influenza A, seasonal coronaviruses, rhinovirus and parainfluenza viruses were detected more frequently (S11 Table). These results should be interpreted cautiously, since the multiplex RT-PCR panels used differ from the point of view of antigens included.

### 3.5. Severe RSV disease

The prespecified outcomes ascribable to severe RSV disease were reported in nine studies [35,41,42,44,50,54,56,63,64–66] (Table 2). Pierangeli et al. [56] reported that 63.1%, 63.3% and 79.3% of adults (both out- and inpatients) aged 16–65, 65–80 and >80 years had antibiotic prescriptions. The study, however, did not reported raw data. In an outpatient cohort (n = 2551) of adults aged ≥60 years [42], none of seven cases of community-acquired pneumonia (CAP) tested positive for RSV. Ciotti et al. [50] reported that among 23 hospitalized cases of viral pneumonia, one (4.3%) was due to RSV. In a multi-season (from 2009/2010 to 2015/2016) study by Piralla et al. [41], the prevalence of RSV among subjects (11/376) with CAP admitted to ICU was 2.9%. Among 19 RSV-positive hematological patients [44], two (10.5%) developed LRTD and the 30-day mortality was 0% (95% CI: 0–17.6%). Spagnolello et al. [54] reported that among CAP patients who referred to emergency departments (n = 75), three tested positive for RSV (4.0%; 95% CI: 1.5–13.1%). However, the study period was limited to one month only (from 15 January to 22 February 2019) [54].

**Table 2.** Severe RSV-related outcomes, by study.

| Study | Age, years | Outcome | % (n/N) | 95% CI |
| --- | --- | --- | --- | --- |
| Cocchio 2023 [35] | 50–69 | % in-hospital mortality | 4.9 (4/81) | 1.4–12.2 |
|  | ≥70 |  | 7.1 (12/169) | 3.7–12.1 |
| Piralla 2017 [41] | ≥18 | % RSV detection in patients with CAP admitted to ICU | 2.9 (11/376) | 1.5–5.2 |
| Ansaldi 2012 [42] | ≥60 | % RSV detection in patients with CAP | 0 (0/7) | 0–41.0 |
| Mikulska 2014 [44] | ≥18 | % RSV-positive hematological patients who developed LRTD | 10.5 (2/19) | 1.3–33.1 |
|  |  | % 30-day mortality | 0 (0/19) | 0–17.6 |
| Ciotti 2020 [50] | ≥18 | % RSV detection in patients hospitalized for viral pneumonia | 4.3 (1/23) | 0.1–21.9 |
| Spagnolello 2021 [54] | >18 | % RSV detection in ED patients with CAP | 4.0 (3/75) | 1.5–13.1 |
| Pierangeli 2022 [56] | 16–65 | % antibiotic use in RSV-positive subjects | 63.1 (NA) | NA |
|  | 65–80 |  | 63.3 (NA) | NA |
|  | >80 |  | 79.3 (NA) | NA |
| Santus 2023 [63] | ≥18 | % antibiotic use in RSV-positive inpatients | 78.7 (48/61) | 66.3–88.1 |
|  |  | Median length of hospital stay, days | 13 <sup>a</sup> | 7–22 <sup>b</sup> |
|  |  | % RSV-positive inpatients with acute respiratory failure, need for respiratory support, shock, sepsis, ICU admission or in-hospital death | 70.5 (43/61) | 57.4–81.5 |
|  |  | % RSV-positive inpatients with ICU admission | 6.6 (4/61) | 1.8–15.9 |
|  |  | % in-hospital mortality | 6.6 (4/61) | 1.8–15.9 |
| Boattini 2021–2023 [64–66] | ≥18 | % pneumonia in hospitalized RSV-positive subjects | 58.1 (25/43) | 42.1–73.0 |
|  | ≥18 | Median length of hospital stay, days | 19 <sup>a</sup> | 11–30 <sup>b</sup> |
|  | ≥65 |  | 23 <sup>a</sup> | 8–34 <sup>b</sup> |
|  | ≥18 | % invasive and non-invasive mechanical ventilation in hospitalized RSV-positive subjects | 30.2 (13/43) | 17.2–46.1 |
|  | ≥65 |  | 27.3 (3/11) | 6.0–61.0 |
|  | ≥18 | % in-hospital mortality | 16.3 (7/43) | 6.8–30.7 |
|  | ≥65 |  | 9.1 (1/11) | 0.2–41.3 |

<sup>a</sup>Median; <sup>b</sup>Interquartile range; CAP, community-acquired pneumonia; CI, Confidence interval; ED, Emergency department; ICU, Intensive care unit; LRTD, Lower respiratory tract disease; NA, Not available.

In a hospitalized cohort of RSV-positive adults (n = 61) [63], severe RSV disease (defined as presence of at least one of the following: acute respiratory failure, need for respiratory support, shock, sepsis, ICU admission or in-hospital death) was observed in 70.5% of patients. Antibiotics, systemic steroids, inhaled steroids, oseltamivir and other antivirals were administered to 79.0%, 65.6%, 70.0%, 13.1% and 8.2% of patients respectively. The median length of stay was 13 (IQR: 7–22) days. ICU admission and in-hospital mortality rates were both 6.6%.

In a retrospective cohort study by Boattini et al. [64–66], 58.1% (n = 43) of RSV-positive hospitalized adults ≥18 years had radiologically-confirmed pneumonia. The median length of stay for the entire cohort was 19 (IQR: 11–30) days, while it increased up to 23 (IQR: 8–34) days for adults aged ≥65 years. A total of 30.2% of subjects needed invasive and non-invasive mechanical ventilation, while the in-hospital mortality was 16.3%. Cocchio et al. [35] analyzed HDRs for RSV-specific ICD-9 codes in Veneto for 15 consecutive years (2007– 2021). Of the total of 6961 (5818, 741 and 402 for acute bronchiolitis due to RSV, pneumonia due to RSV and RSV, respectively) hospitalizations, 81 (1.2%; 8, 41 and 32 for acute bronchiolitis due to RSV, pneumonia due to RSV and RSV, respectively) and 169 (2.4%; 31, 74 and 64 for acute bronchiolitis due to RSV, pneumonia due to RSV and RSV, respectively) encounters occurred in adults aged 50–69 and ≥70 years, respectively. The length of stay increased linearly with the increasing age for acute bronchiolitis due to RSV (β = 0.080; 95% CI: 0.059–0.100; P < 0.001), pneumonia due to RSV (β = 0.109; 95% CI: 0.083–0.135; P < 0.001) and RSV (β = 0.063; 95% CI: 0.045–0.081; P < 0.001), respectively. Of 23 deaths identified, 73.9% (17/23) were registered in subjects aged ≥50 years. The overall in-hospital mortality rate was 4.9% (4/81; 0%, 7.3% and 3.1% for acute bronchiolitis due to RSV, pneumonia due to RSV and RSV, respectively) and 7.1% (12/169; 12.9%, 10.8% and 0% for acute bronchiolitis due to RSV, pneumonia due to RSV and RSV, respectively) in subjects aged 50–69 and ≥70 years, respectively.

As reported in Table 2, only the outcome of in-hospital mortality was reported in more studies [35,63–66]. In a pooled analysis of four estimates (S7 Fig), the in-hospital mortality was 7.2% (95% CI: 4.7–10.3%). The heterogeneity was relatively low (I^2^ = 30.6%) and the 95% PI was 2.2–14.5%.

## 4. Discussion

This SRMA showed that RSV is a frequent respiratory pathogen that poses a measurable burden to Italian adults. However, this BoD is unevenly distributed within the Italian adult population, being higher in older age groups and subjects with underlying health conditions. Our findings may inform principal stakeholders and support policy makers on the introduction of the recently authorized adults RSV vaccines [13] with the aim to ensure effective and coherent resource allocation. Analogously, our study is a good starting point for the upcoming cost-effectiveness and budget impact models. We finally identified principal data gaps, which must be addressed by future research.

Our meta-analysis showed that 4.5% of respiratory samples tests positive for RSV and the 95% PI suggested that the next estimate would be between 0% and 16%. This uncertainty reflects both natural between-season variation in the circulation of RSV and characteristics of the population surveilled. We indeed showed that compared with hospitalized individuals, community-dwelling adults had higher positivity prevalence (4.9% vs 2.9%). At the same time, compared with working-age adults (3.5%), RSV prevalence was higher in older adults (4.4%) and individuals with immunosuppressive disorders (11.5%). It should be stressed that all these pooled estimates are probably conservative, as a certain level of underestimation is likely for several reasons. First, most primary care studies were conducted within the existing influenza surveillance framework that usually relied on ILI-based case definitions. Different studies [53,67,68] have proved that a significant proportion of RSV-positive adults have no fever and therefore ILI and fever-based case definitions intrinsically underestimate the true incidence of RSV. Second, several hospital-based studies considered only one type of specimen. It has been, for example, shown [69] that adding RT-PCR of sputum to that of nasal or nasopharyngeal swab specimens increases RSV detection by 39–100%. Indeed, RSV may migrate from the upper to lower respiratory tracts and thus RT-PCR performed on naso/oropharyngeal swabs would produce a false-negative result [28].

When comparing our results to those of the global-level SRMAs [5–8], we noted both similarities and differences. Nguyen-Van-Tam et al. [7] have reported that the cumulative attack rate of RSV in older adults ranged from 0.27‰ to 108‰. This 400-fold difference [7] is unlikely to be explained by the epidemiology of RSV alone, but is rather linked to the design of single studies. As for RSV incidence in industrialized countries, Shi et al. obtained pooled estimates of 6.7‰ (95% CI: 1.4–31.5‰) [5] and 30.3‰ (95% CI: 5.3–59.9‰) [8] for older and high-risk adults, respectively. In Italy, the available incidence estimates were 1.9 ‰ [42] and 4.4– 10.9% [44,57] for older adults and adults with immunosuppressive conditions, respectively. Italian data, however, came from single-season studies and thus affected by RSV circulation patterns in those particular seasons. RSV positivity prevalence appears instead more consistent. For instance, a meta-analysis of European studies by Tin Tin Htar et al. [6] reported that 7% (95% CI: 4–11%), 9% (95% CI: 4–17%) and 10% (95% CI: 5–16%) of ILI/ARI cases in all adults, subjects aged <50 and ≥50 years, respectively, tested positive for RSV. Shi et al. [5] estimated that RSV positivity among hospitalized older adults with ARI was 4.4% (95% CI: 3.0–6.5%). Our meta-analytical proportions were in line with these figures. For what concerns mortality indicators, the pooled in-hospital mortality in Italy was 7.2%. In comparison, the case-fatality proportions ranged from 1.6% (95% CI: 0.7–3.8%) [5] to 8.2% (95% CI 5.5–11.9%) [7] in the available global-kevel SRMAs. Apart from age, underlying immunosuppressive conditions and setting, RSV positivity prevalence was associated with the study location and sample size. Studies conducted in the north of Italy reported on average a higher frequency of RSV. Compared with central and southern regions, mean winter temperature in northern regions is lower [70]. Lower ambient temperature is a well-known positive predictor of RSV detection frequency [71–73]. Smaller studies tended to report a higher RSV prevalence, which is consistent with the small study effect. Fortunately, the double arcsine transformation prevents the undue large weights for these studies [74]. Future epidemiological studies should *a priori* determine an adequate number of subjects to be tested for RSV. Despite a relatively high number of publications identified, most studies were limited to quantifying prevalence of RSV following clinical request or in the context of influenza surveillance and were not specifically designed to investigate RSV epidemiology and the associated BoD. Only a few papers went beyond mere positivity prevalence reports. Currently, no cohort studies have evaluated the natural history of RSV disease in Italian adults. In particular, it remains unclear how many RSV-positive adult outpatients develop complications and being hospitalized. Some of these data gaps could be filled by a retrospective analysis of HDRs, but only if matched with laboratory data. Indeed, many RSV-positive patients could be attributed alternative diagnostic codes. Despite being highly specific and useful for the analysis of case-fatality rates, resource use and temporal trends, studies of RSV-specific HDRs in adults inevitably lead to the underestimation of RSV BoD. Cai et al. [24] have documented that the RSV-specific ICD-10 codes had a very low sensitivity of 6%. Retrospective cohort studies based on HDRs only (i.e., without matching with laboratory data) would identify an implausibly low number of RSV-specific diagnostic codes [35,75]. Another important data gap concerns RSV epidemiology and BoD in residents of long-term care facilities, who may be the primary target population for vaccination.

As we discussed earlier, the main limitations of the current evidence, which may underestimate RSV BoD, include the predominant use of influenza-specific case definitions, reliance on a single specimen type, lack of studies on the natural history of an RSV episode and some important BoD indicators like hospitalization rate.

At review level, we must acknowledge that some potentially relevant studies were not identified because their primary goal was not related to RSV and thus were indexed alternatively.

The approval of the new vaccines depends on national authorities. This decision occurs within the framework of developing national immunization programs and necessitates the utilization of assessment tools, such as the health technology assessment (HTA) [76]. This process relies on a comprehensive understanding of the epidemiology, BoD and pertinent economic analyses. Furthermore, HTA organizations are also evaluating the implementation of guidelines that take into account, in the assessment of new technologies, also their societal value [77]. This objective aligns with the "Broader value of vaccines" framework proposed by Bell et al. [78]. The framework encompasses various vaccine effects, including strictly health-related impacts, focusing on the health of vaccinated individuals; general health effects, considering the influence of vaccination on the health of the unvaccinated population; economic effects on the healthcare system, involving the costs and corresponding budgetary compensations; and societal economic effects, evaluating broader economic of vaccines impacts such as effects on productivity and macroeconomic growth from a societal perspective.

Incorporating these diverse effects is crucial in the HTA processes [77]. Enhancing the vaccines HTA involves considering several additional factors, including broader cost offsets within the healthcare system, the influence of vaccination on the quality of life of caregivers, its effectiveness in reducing viral transmission, prevention of antimicrobial resistance, and the macroeconomic impacts of vaccination [79]. Moreover, there is a requirement for novel economic models capable of capturing not only the simple cost/benefit ratio of vaccination but also its broader value [80].

As part of future research and in light of the availability of new vaccines, it is evident that the generation of new tools, evidence, and data is imperative to delve deeper into the long-term benefits arising from vaccination, explore macroeconomic implications, understand community impact, and address issues pertaining to equity, accessibility, and appropriate utilization. Moreover, it is crucial to emphasize greater involvement of citizens in the implementation of vaccination decisions [81].

In conclusion, this article provides a comprehensive assessment of the RSV epidemiology and its BoD in the Italian adult population. A systematic assessment of the full value of a new vaccination necessitates access to national data regarding the epidemiological burden of the disease and the presence of effective surveillance systems. Data plays a pivotal role in generating knowledge and evidence about diseases. Only through evidence can priority actions for the control of infectious diseases be discerned, facilitating the promotion of an informed decision-making process. This, in turn, supports the implementation of value-based immunization strategies. In this paper, we comprehensively assessed RSV epidemiology and BoD in the Italian adult population. Within its shortcomings, this SRMA supports the need to prioritize evaluation of the novel RSV vaccines by Italian decision makers, especially for older adults and those affected by immunosuppressive conditions.

## Data availability statement

All raw data are within the manuscript and associated supporting materials. Further details may be obtained from the corresponding author.

## Funding

This research received no funding.

## Competing interests

AD and GEC declare no conflicts of interest related to this work.

## Supporting information captions

**S1 Fig. RSV positivity prevalence among Italian adults of any age, by study period in relation to the COVID-19 pandemic.**

**S2 Fig. RSV positivity prevalence among Italian adults of any age, by study geographic area**

**S3 Fig. RSV positivity prevalence among Italian adults of any age, by study sample size.**

**S4 Fig. Prevalence of RSV subtype B among Italian adults of any age (prevalence of RSV subtype A may be computed as 1 – prevalence of RSV B).**

**S5 Fig. Frequency of viral co-detections among RSV-positive Italian adults of any age.**

**S6 Fig. In-hospital mortality among RSV-positive Italian adults of any age.**

**S1 Table. PRISMA (Preferred Reporting Items for Systematic reviews and Meta-Analyses) checklist.**

**S2 Table. Algorithm for the automatic search, by citation database.**

**S3 Table. Excluded studies with reasons.**

**S4 Table. Syndromic definitions used the studies analyzed.**

**S5 Table. Risk of bias of the studies included according to the Joanna Briggs Institute (JBI) checklist for prevalence/incidence studies.**

**S6 Table. RSV positivity prevalence among Italian adults of any age, by setting.**

**S7 Table. RSV positivity prevalence among Italian working-age adults, by setting. S8 Table. RSV positivity prevalence among Italian older adults, by setting.**

**S9 Table. Meta-regression analysis to investigate sources of heterogeneity in RSV positivity prevalence among Italian adults of any age.**

**S10 Table. RSV positivity prevalence among Italian adults, by subtype.**

**S11 Table. Frequency of viral co-detections among RSV-positive Italian adults of any age.**

## Notes

### Competing Interest Statement

The authors have declared no competing interest.

### Funding Statement

The author(s) received no specific funding for this work.

## References

1. McLaughlin JM, Khan F, Begier E, Swerdlow DL, Jodar L, Falsey AR. Rates of medically attended RSV among US adults: A systematic review and meta-analysis. Open Forum Infect Dis. 2022;9(7):ofac300. doi: 10.1093/ofid/ofac300.

2. Rafferty E, Paulden M, Buchan SA, Robinson JL, Bettinger JA, Kumar M, et al. Evaluating the individual healthcare costs and burden of disease associated with RSV across age groups. Pharmacoeconomics. 2022;40(6):633–645. doi: 10.1007/s40273-022-01142-w.

3. Osei-Yeboah R, Spreeuwenberg P, Del Riccio M, Fischer TK, Egeskov-Cavling AM, Bøås H, et al. Estimation of the number of RSV-associated hospitalisations in adults in the European Union. J Infect Dis. 2023;228(11):1539–1548. doi: 10.1093/infdis/jiad189.

4. Du Y, Yan R, Wu X, Zhang X, Chen C, Jiang D, et al. Global burden and trends of respiratory syncytial virus infection across different age groups from 1990 to 2019: A systematic analysis of the Global Burden of Disease 2019 Study. Int J Infect Dis. 2023;135:70–76. doi: 10.1016/j.ijid.2023.08.008.

5. Shi T, Denouel A, Tietjen AK, Campbell I, Moran E, Li X, et al. Global disease burden estimates of respiratory syncytial virus-associated acute respiratory infection in older adults in 2015: A systematic review and meta-analysis. J Infect Dis. 2020;222(Suppl 7):S577–S583. doi: 10.1093/infdis/jiz059.

6. Tin Tin Htar M, Yerramalla MS, Moïsi JC, Swerdlow DL. The burden of respiratory syncytial virus in adults: a systematic review and meta-analysis. Epidemiol Infect. 2020;148:e48. doi: 10.1017/S0950268820000400.

7. Nguyen-Van-Tam JS, O’Leary M, Martin ET, Heijnen E, Callendret B, Fleischhackl R, et al. Burden of respiratory syncytial virus infection in older and high-risk adults: a systematic review and meta-analysis of the evidence from developed countries. Eur Respir Rev. 2022;31(166):220105. doi: 10.1183/16000617.0105-2022.

8. Shi T, Vennard S, Jasiewicz F, Brogden R, Nair H; RESCEU Investigators. Disease burden estimates of respiratory syncytial virus related acute respiratory infections in adults with comorbidity: A systematic review and meta-analysis. J Infect Dis. 2022;226(Suppl 1):S17–S21. doi: 10.1093/infdis/jiab040.

9. Maggi S, Veronese N, Burgio M, Cammarata G, Ciuppa ME, Ciriminna S, et al. Rate of hospitalizations and mortality of respiratory syncytial virus infection compared to influenza in older people: A systematic review and meta-analysis. Vaccines (Basel). 2022;10(12):2092. doi: 10.3390/vaccines10122092.

10. Li Y, Kulkarni D, Begier E, Wahi-Singh P, Wahi-Singh B, Gessner B, et al. Adjusting for case under-ascertainment in estimating RSV hospitalisation burden of older adults in high-income countries: A systematic review and modelling study. Infect Dis Ther. 2023;12(4):1137–1149. doi: 10.1007/s40121-023-00792-3.

11. Haynes AK, Manangan AP, Iwane MK, Sturm-Ramirez K, Homaira N, Brooks WA, et al. Respiratory syncytial virus circulation in seven countries with Global Disease Detection Regional Centers. J Infect Dis. 2013;208(Suppl 3):S246–S254. doi: 10.1093/infdis/jit515.

12. Staadegaard L, Caini S, Wangchuk S, Thapa B, de Almeida WAF, de Carvalho FC, et al. The global epidemiology of RSV in community and hospitalized care: Findings from 15 countries. Open Forum Infect Dis. 2021;8(7):ofab159. doi: 10.1093/ofid/ofab159.

13. Kotton CN. More protection against respiratory viral infection: Respiratory syncytial virus vaccines for adults aged 60 years and older. Ann Intern Med. 2023;176(10):1419–1421. doi: 10.7326/M23-2196.

14. Centers for Disease Control and Prevention (CDC). Healthcare providers: RSV vaccination for adults 60 years of age and over. Available at: https://www.cdc.gov/vaccines/vpd/rsv/hcp/older-adults.html#:~:text=CDC%20recommends%20that%20adults%2060,RSV%20vaccination%20will%20be%20beneficial.

15. United Kingdom Department of Health & Social Care. Respiratory syncytial virus (RSV) immunisation programme for infants and older adults: JCVI full statement, 11 September 2023. Available at: https://www.gov.uk/government/publications/rsv-immunisation-programme-jcvi-advice-7-june-2023/respiratory-syncytial-virus-rsv-immunisation-programme-for-infants-and-older-adults-jcvi-full-statement-11-september-2023.

16. Levine MM, Levine OS. Influence of disease burden, public perception, and other factors on new vaccine development, implementation, and continued use. Lancet. 1997;350(9088):1386–1392. doi: 10.1016/S0140-6736(97)03253-4.

17. Munn Z, Stern C, Aromataris E, Lockwood C, Jordan Z. What kind of systematic review should I conduct? A proposed typology and guidance for systematic reviewers in the medical and health sciences. BMC Med Res Methodol. 2018;18(1):5. doi: 10.1186/s12874-017-0468-4.

18. Barker TH, Migliavaca CB, Stein C, Colpani V, Falavigna M, Aromataris E, et al. Conducting proportional meta-analysis in different types of systematic reviews: A guide for synthesisers of evidence. BMC Med Res Methodol. 2021;21(1):189. doi: 10.1186/s12874-021-01381-z.

19. Boccalini S, Bonito B, Salvati C, Del Riccio M, Stancanelli E, Bruschi M, et al. Human respiratory syncytial virus epidemiological burden in pediatric outpatients in Italy: A systematic review. Vaccines (Basel). 2023;11(9):1484. doi: 10.3390/vaccines11091484.

20. Page MJ, McKenzie JE, Bossuyt PM, Boutron I, Hoffmann TC, Mulrow CD, et al. The PRISMA 2020 statement: an updated guideline for reporting systematic reviews. BMJ. 2021;372:n71. doi: 10.1136/bmj.n71.

21. Munn Z, Moola S, Lisy K, Riitano D, Tufanaru C. Methodological guidance for systematic reviews of observational epidemiological studies reporting prevalence and cumulative incidence data. Int J Evid Based Healthc. 2015;13(3):147–53. doi: 10.1097/XEB.0000000000000054.

22. Domnich A, Calabrò GE. Burden of RSV in Italian adults: Protocol for a systematic review and meta-analysis. Available at: https://www.protocols.io/view/burden-of-rsv-in-italian-adults-protocol-for-a-sys-5qpvo32odv4o/v1.

23. Pisesky A, Benchimol EI, Wong CA, Hui C, Crowe M, Belair MA, et al. Incidence of hospitalization for respiratory syncytial virus infection amongst children in Ontario, Canada: A population-based study using validated health administrative data. PLoS One. 2016;11(3):e0150416. doi: 10.1371/journal.pone.0150416.

24. Cai W, Tolksdorf K, Hirve S, Schuler E, Zhang W, Haas W, et al. Evaluation of using ICD-10 code data for respiratory syncytial virus surveillance. Influenza Other Respir Viruses. 2020;14(6):630–637. doi: 10.1111/irv.12665.

25. Haddaway NR, Collins AM, Coughlin D, Kirk S. The role of Google Scholar in evidence reviews and its applicability to grey literature searching. PLoS One. 2015;10(9):e0138237. doi: 10.1371/journal.pone.0138237.

26. Hamid S, Winn A, Parikh R, Jones JM, McMorrow M, Prill MM, et al. Seasonality of respiratory syncytial virus - United States, 2017-2023. MMWR Morb Mortal Wkly Rep. 2023;72(14):355–361. doi: 10.15585/mmwr.mm7214a1.

27. Bergeri I, Whelan MG, Ware H, Subissi L, Nardone A, Lewis HC, et al. Global SARS-CoV-2 seroprevalence from January 2020 to April 2022: A systematic review and meta-analysis of standardized population-based studies. PLoS Med. 2022;19(11):e1004107. doi: 10.1371/journal.pmed.1004107.

28. Rozenbaum MH, Begier E, Kurosky SK, Whelan J, Bem D, Pouwels KB, et al. Incidence of respiratory syncytial virus infection in older adults: Limitations of current data. Infect Dis Ther. 2023;12(6):1487– 1504. doi: 10.1007/s40121-023-00802-4.

29. The Cochrane Collaboration. Cochrane handbook for systematic reviews of interventions, version 5.1.0, updated March 2011. Meta-regression. Available at: https://handbook-5-1.cochrane.org/chapter_9/9_6_4_meta_regression.htm#:~:text=Meta%2Dregression%20should%20generally%20not,one%20or%20more%20explanatory%20variables.

30. Rovida F, Percivalle E, Zavattoni M, Torsellini M, Sarasini A, Campanini G, et al. Monoclonal antibodies versus reverse transcription-PCR for detection of respiratory viruses in a patient population with respiratory tract infections admitted to hospital. J Med Virol. 2005;75(2):336–347. doi: 10.1002/jmv.20276.

31. Minosse C, Selleri M, Zaniratti MS, Cappiello G, Longo R, Schifano E, et al. Frequency of detection of respiratory viruses in the lower respiratory tract of hospitalized adults. J Clin Virol. 2008;42(2):215–220. doi: 10.1016/j.jcv.2008.01.012.

32. Puzelli S, Valdarchi C, Ciotti M, Dorrucci M, Farchi F, Babakir-Mina M, et al. Viral causes of influenza-like illness: Insight from a study during the winters 2004-2007. J Med Virol. 2009;81(12):2066–71. doi: 10.1002/jmv.21610.

33. Costa C, Libertucci D, Solidoro P, Sinesi F, Bergallo M, Margio S, et al. Rapid shell vial culture for the detection of respiratory viruses from bronchoalveolar lavage in immunocompromised patients. Panminerva Med. 2007;49(1):1–6.

34. Gerna G, Piralla A, Rovida F, Rognoni V, Marchi A, Locatelli F, et al. Correlation of rhinovirus load in the respiratory tract and clinical symptoms in hospitalized immunocompetent and immunocompromised patients. J Med Virol. 2009;81(8):1498–1507.

35. Cocchio S, Prandi GM, Furlan P, Venturato G, Saia M, Marcon T, et al. Respiratory syncytial virus in Veneto Region: Analysis of hospital discharge records from 2007 to 2021. Int J Environ Res Public Health. 2023;20(5):4565. doi: 10.3390/ijerph20054565.

36. Gambarino S, Mantovani S, Astegiano S, Libertucci D, Solidoro P, Baldi S, et al. Lower respiratory tract viral infections in hospitalized adult patients. Minerva Med. 2009;100(5):349–355.

37. Paba P, Farchi F, Mortati E, Ciccozzi M, Piperno M, Perno CF, et al. Screening of respiratory pathogens by Respiratory Multi Well System (MWS) r-gene™ assay in hospitalized patients. New Microbiol. 2014;37(2):231–236.

38. Pierangeli A, Scagnolari C, Selvaggi C, Verzaro S, Spina MT, Bresciani E, et al. Rhinovirus frequently detected in elderly adults attending an emergency department. J Med Virol. 2011;83(11):2043–2047. doi: 10.1002/jmv.22205.

39. Nisii C, Meschi S, Selleri M, Bordi L, Castilletti C, Valli MB, et al. Frequency of detection of upper respiratory tract viruses in patients tested for pandemic H1N1/09 viral infection. J Clin Microbiol. 2010;48(9):3383–3385. doi: 10.1128/JCM.01179-10.

40. Tramuto F, Maida CM, Napoli G, Mammina C, Casuccio A, Cala’ C, et al. Burden and viral aetiology of influenza-like illness and acute respiratory infection in intensive care units. Microbes Infect. 2016;18(4):270–276. doi: 10.1016/j.micinf.2015.11.008.

41. Piralla A, Mariani B, Rovida F, Baldanti F. Frequency of respiratory viruses among patients admitted to 26 Intensive Care Units in seven consecutive winter-spring seasons (2009-2016) in Northern Italy. J Clin Virol. 2017;92:48–51. doi: 10.1016/j.jcv.2017.05.004.

42. Ansaldi F, De Florentiis D, Parodi V, Rappazzo E, Coppelli M, Martini M, et al. Bacterial carriage and respiratory tract infections in subjects > or = 60 years during an influenza season: implications for the epidemiology of community acquired pneumonia and influenza vaccine effectiveness. J Prev Med Hyg. 2012;53(2):94–97.

43. Bigliardi S, Morselli M, Potenza L, Riva G, Coluccio V, Maccaferri M, et al. Epidemiology and clinical outcome of lower respiratory tract infections by respiratory syncytial virus or parainfluenza virus type 3 in adults receiving treatment for either acute leukemia or severe aplastic anemia: A retrospective single center study. Ann Hematol. 2015;94(11):1931–1934. 10.1007/s00277-015-2455-3.

44. Mikulska M, Del Bono V, Gandolfo N, Dini S, Dominietto A, Di Grazia C, et al. Epidemiology of viral respiratory tract infections in an outpatient haematology facility. Ann Hematol. 2014;93(4):669–676. doi: 10.1007/s00277-013-1912-0.

45. Passi A, Pagani C, Gramegna D, Daffini R, Pollara CP, Signorini L, et al. Respiratory viruses are a significant clinical problem in haematological patients with underestimated adverse outcome: A single institution 9-years experience. Haematologica 2019;104(S2):29–30.

46. Pellegrinelli L, Galli C, Bubba L, Cereda D, Anselmi G, Binda S, et al. Respiratory syncytial virus in influenza-like illness cases: Epidemiology and molecular analyses of four consecutive winter seasons (2014-2015/2017-2018) in Lombardy (Northern Italy). J Med Virol. 2020;92(12):2999–3006. doi: 10.1002/jmv.25917.

47. Costa E, Giberti I, Scarpaleggia M, Massolo L, Massaro E, Trombetta CS, et al. Retrospective analysis of viral diagnostics data of patients referred to the Policlinico San Martino Hospital in Genoa with acute respiratory infection during eight sequential seasons (2014–2022). Presented at the 33rd European Congress of Clinical Microbiology and Infectious Diseases (ECCMID), Copenhagen (Denmark), 15–18 April 2023.

48. Tramuto F, Maida CM, Di Naro D, Randazzo G, Vitale F, Restivo V, et al. Respiratory syncytial virus: New challenges for molecular epidemiology surveillance and vaccination strategy in patients with ILI/SARI. Vaccines (Basel). 2021;9(11):1334. doi: 10.3390/vaccines9111334.

49. Leli C, Di Matteo L, Gotta F, Vay D, Piceghello A, Cornaglia E, et al. Prevalence of respiratory viruses by Multiplex PCR: a four-and-a-half year retrospective study in an Italian general hospital. Infez Med. 2021;29(1):94–101.

50. Ciotti M, Maurici M, Santoro V, Coppola L, Sarmati L, De Carolis G, et al. Viruses of respiratory tract: an observational retrospective study on hospitalized patients in Rome, Italy. Microorganisms. 2020;8(4):501. doi: 10.3390/microorganisms8040501.

51. De Francesco MA, Pollara C, Gargiulo F, Giacomelli M, Caruso A. Circulation of respiratory viruses in hospitalized adults before and during the COVID-19 pandemic in Brescia, Italy: A retrospective study. Int J Environ Res Public Health. 2021;18(18):9525. doi: 10.3390/ijerph18189525.

52. Galli C, Pellegrinelli L, Del Castillo G, Forni G, Gandolfi CE, Mosillo M, et al. Self-sampling of oropharyngeal swabs among healthcare workers for molecular detection of respiratory viruses: A valuable approach for epidemiological studies and surveillance programs. Front Public Health. 2020;8:511669. doi: 10.3389/fpubh.2020.511669.

53. Domnich A, Orsi A, Ogliastro M, Ferrari A, Bruzzone B, Panatto D, Icardi G. Influenza-like illness surveillance may underestimate the incidence of respiratory syncytial virus in adult outpatients. Int J Infect Dis. Forthcoming.

54. Spagnolello O, Pierangeli A, Cedrone MC, Di Biagio V, Gentile M, Leonardi A, et al. Viral community acquired pneumonia at the emergency department: Report from the pre COVID-19 age. J Med Virol. 2021;93(7):4399–4404. doi: 10.1002/jmv.26980.

55. Sberna G, Lalle E, Valli MB, Bordi L, Garbuglia AR, Amendola A. Changes in the Circulation of Common Respiratory Pathogens among Hospitalized Patients with Influenza-like Illnesses in the Lazio Region (Italy) during Fall Season of the Past Three Years. Int J Environ Res Public Health. 2022;19(10):5962. doi: 10.3390/ijerph19105962.

56. Pierangeli A, Piralla A, Uceda Renteria S, Giacomel G, Lunghi G, Pagani E, et al. Multicenter epidemiological investigation and genetic characterization of respiratory syncytial virus and metapneumovirus infections in the pre-pandemic 2018-2019 season in northern and central Italy. Clin Exp Med. 2023;23(6):2725–2737. doi: 10.1007/s10238-022-00973-3.

57. Scagnolari C, Bitossi C, Frasca F, Viscido A, Oliveto G, Scordio M, et al. No detection of SARS-CoV-2 in cystic fibrosis patients at the Regional (Lazio) Reference Center for CF in Italy. J Cyst Fibros. 2020;19(5):837–838. doi: 10.1016/j.jcf.2020.06.018.

58. Galli C, Pellegrinelli L, Bubba L, Primache V, Anselmi G, Delbue S, et al. When the COVID-19 pandemic surges during influenza season: Lessons learnt from the sentinel laboratory-based surveillance of influenza-like illness in Lombardy during the 2019-2020 season. Viruses. 2021;13(4):695. doi: 10.3390/v13040695.

59. Treggiari D, Piubelli C, Formenti F, Silva R, Perandin F. Resurgence of respiratory virus after relaxation of COVID-19 containment measures: A real-world data study from a regional hospital of Italy. Int J Microbiol. 2022;2022:4915678. doi: 10.1155/2022/4915678.

60. Calderaro A, De Conto F, Buttrini M, Piccolo G, Montecchini S, Maccari C, et al. Human respiratory viruses, including SARS-CoV-2, circulating in the winter season 2019-2020 in Parma, Northern Italy. Int J Infect Dis. 2021;102:79–84. doi: 10.1016/j.ijid.2020.09.1473.

61. Milano G, Capitani E, Camarri A, Bova G, Capecchi PL, Lazzeri G, et al. Surveillance of influenza and other airborne transmission viruses during the 2021/2022 season in hospitalized subjects in Tuscany, Italy. Vaccines (Basel). 2023;11(4):776. doi: 10.3390/vaccines11040776.

62. Panatto D, Domnich A, Lai PL, Ogliastro M, Bruzzone B, Galli C, et al. Epidemiology and molecular characteristics of respiratory syncytial virus (RSV) among italian community-dwelling adults, 2021/22 season. BMC Infect Dis. 2023;23(1):134. doi: 10.1186/s12879-023-08100-7.

63. Santus P, Radovanovic D, Gismondo MR, Rimoldi SG, Lombardi A, Danzo F, et al. Respiratory syncytial virus burden and risk factors for severe disease in patients presenting to the emergency department with flu-like symptoms or acute respiratory failure. Respir Med. 2023;218:107404. doi: 10.1016/j.rmed.2023.107404

64. Boattini M, Almeida A, Christaki E, Marques TM, Tosatto V, Bianco G, et al. Severity of RSV infection in Southern European elderly patients during two consecutive winter seasons (2017-2018). J Med Virol. 2021;93(8):5152–5157. doi: 10.1002/jmv.26938.

65. Almeida A, Boattini M, Christaki E, Moreira Marques T, Moreira I, Cruz L, et al. Comparative virulence of seasonal viruses responsible for lower respiratory tract infections: a southern European multi-centre cohort study of hospital admissions. Infection. 2021;49(3):483–490. doi: 10.1007/s15010-020-01569-3.

66. Boattini M, Charrier L, Almeida A, Christaki E, Moreira Marques T, Tosatto V, et al. Burden of primary influenza and respiratory syncytial virus pneumonia in hospitalised adults: insights from a 2-year multi-centre cohort study (2017-2018). Intern Med J. 2023;53(3):404–408. doi: 10.1111/imj.15583.

67. Sáez-López E, Pechirra P, Costa I, Cristóvão P, Conde P, Machado A, et al. Performance of surveillance case definitions for respiratory syncytial virus infections through the sentinel influenza surveillance system, Portugal, 2010 to 2018. Euro Surveill. 2019;24:1900140. doi: 10.2807/1560-7917.ES.2019.24.45.1900140.

68. Korsten K, Adriaenssens N, Coenen S, Butler CC, Verheij TJM, Bont LJ, et al. World Health Organization influenza-like illness underestimates the burden of respiratory syncytial virus infection in community-dwelling older adults. J Infect Dis. 2022;226:S71–S78. doi: 10.1093/infdis/jiab452.

69. McLaughlin JM, Khan F, Begier E, Swerdlow DL, Jodar L, Falsey AR. Rates of medically attended RSV among US adults: A systematic review and meta-analysis. Open Forum Infect Dis. 2022;9(7):ofac300. doi: 10.1093/ofid/ofac300.

70. Fallani E, Orsi A, Signori A, Icardi G, Domnich A. An exploratory study to assess patterns of influenza- and pneumonia-related mortality among the Italian elderly. Hum Vaccin Immunother. 2021;17(12):5514–5521. doi: 10.1080/21645515.2021.2005381.

71. Noyola DE, Mandeville PB. Effect of climatological factors on respiratory syncytial virus epidemics. Epidemiol Infect. 2008;136(10):1328–1332. doi: 10.1017/S0950268807000143.

72. Lee MH, Mailepessov D, Yahya K, Loo LH, Maiwald M, Aik J. Air quality, meteorological variability and pediatric respiratory syncytial virus infections in Singapore. Sci Rep. 2023;13(1):1001. doi: 10.1038/s41598-022-26184-0.

73. Radhakrishnan D, Ouedraogo A, Shariff SZ, McNally JD, Benchimol EI, Clemens KK. The association between climate, geography and respiratory syncitial virus hospitalizations among children in Ontario, Canada: a population-based study. BMC Infect Dis. 2020;20(1):157. doi: 10.1186/s12879-020-4882-6.

74. Barendregt JJ, Doi SA, Lee YY, Norman RE, Vos T. Meta-analysis of prevalence. J Epidemiol Community Health. 2013;67(11):974–978. doi: 10.1136/jech-2013-203104.

75. Amodio E, Vitale F, d’Angela D, Carrieri C, Polistena B, Spandonaro F, et al. Increased risk of hospitalization for pneumonia in Italian adults from 2010 to 2019: Scientific evidence for a call to action. Vaccines (Basel). 2023;11(1):187. doi: 10.3390/vaccines11010187.

76. O’Rourke B, Oortwijn W, Schuller T; International Joint Task Group. The new definition of health technology assessment: A milestone in international collaboration. Int J Technol Assess Health Care. 2020;36(3):187–190. doi: 10.1017/S0266462320000215

77. Breslau RM, Cohen JT, Diaz J, Malcolm B, Neumann PJ. A review of HTA guidelines on societal and novel value elements. Int J Technol Assess Health Care. 2023;39(1):e31. doi: 10.1017/S026646232300017X.

78. Bell E, Neri M, Steuten L. Towards a broader assessment of value in vaccines: The BRAVE way forward. Appl Health Econ Health Policy. 2022;20(1):105–117. doi: 10.1007/s40258-021-00683-z.

79. Calabro’ GE, Carini E, Tognetto A, Giacchetta I, Bonanno E, Mariani M, et al. The value(s) of vaccination: Building the scientific evidence according to a value-based healthcare approach. Front Public Health. 2022;10:786662. doi: 10.3389/fpubh.2022.786662.

80. Beck E, Biundo E, Devlin N, Doherty TM, Garcia-Ruiz AJ, Postma M, et al. Capturing the value of vaccination within health technology assessment and health economics: Literature review and novel conceptual framework. Vaccine. 2022;40(30):4008–4016. doi: 10.1016/j.vaccine.2022.04.050.

81. de Waure C, Calabrò GE, Ricciardi W; Value(s) of Vaccination Project Steering Committee. Recommendations to drive a value-based decision-making on vaccination. Expert Rev Vaccines. 2022;21(3):289–296. doi: 10.1080/14760584.2022.2021880.

