## Supplementary Figure 1 for "Epidemiology and burden of respiratory syncytial virus in Italian adults: A systematic review and meta-analysis"

**S1 Fig.** RSV positivity prevalence among Italian adults of any age, by study period in relation to the COVID-19 pandemic.

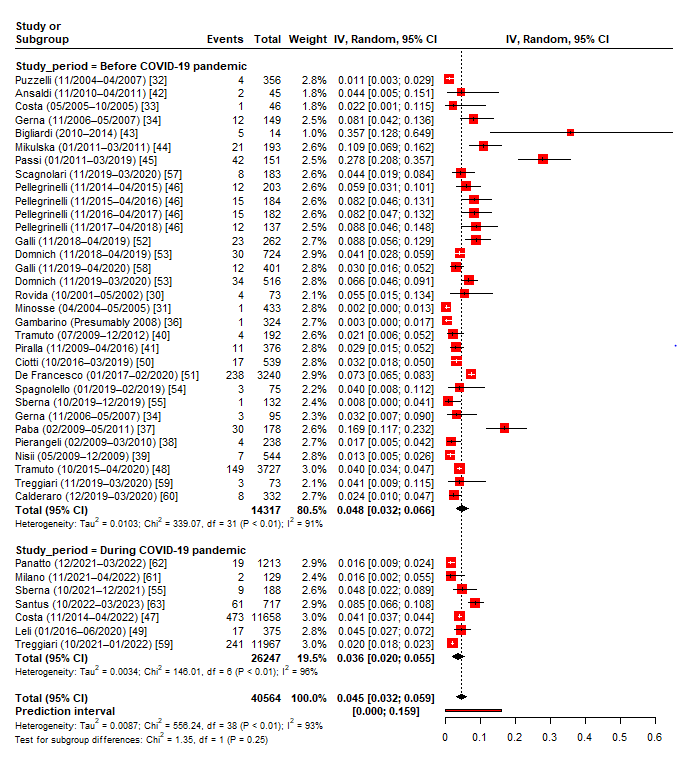
