## Supplementary figures and images for "Epidemiology and burden of respiratory syncytial virus in Italian adults: A systematic review and meta-analysis"

### Supplementary Figure 2

**S2 Fig.** RSV positivity prevalence among Italian adults of any age, by study geographic area.

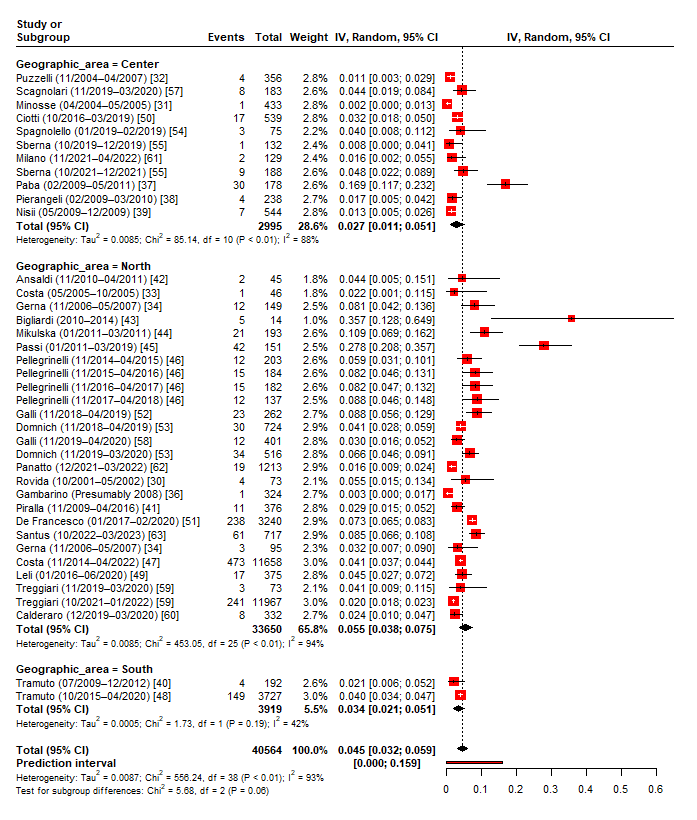

### Supplementary Figure 3

**S3 Fig.** RSV positivity prevalence among Italian adults of any age, by study sample size.

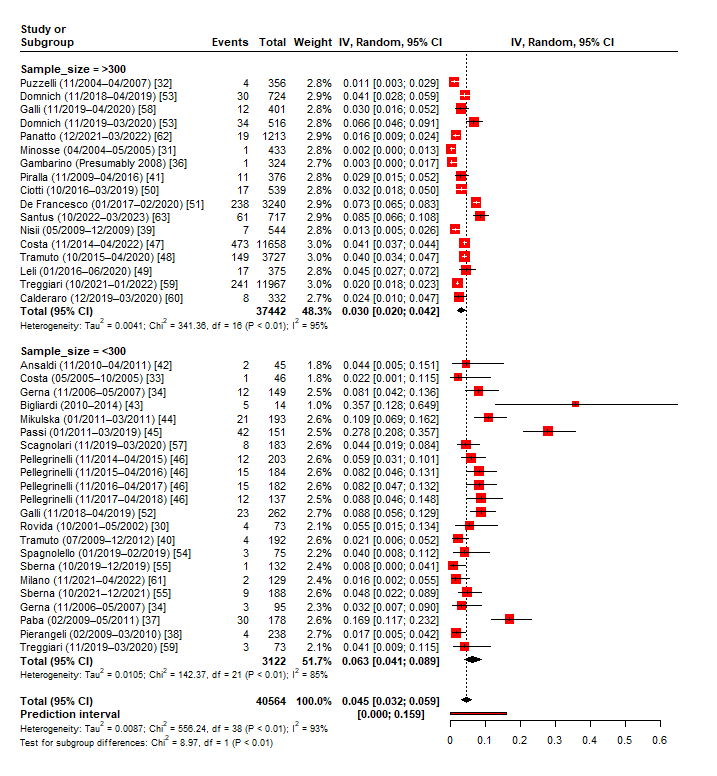

### Supplementary Figure 5

**S5 Fig.** Frequency of viral co-detections among RSV-positive Italian adults of any age.

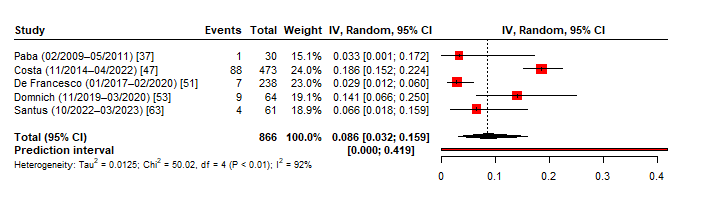

### Supplementary Figure 6

**S6 Fig.** In-hospital mortality among RSV-positive Italian adults of any age.

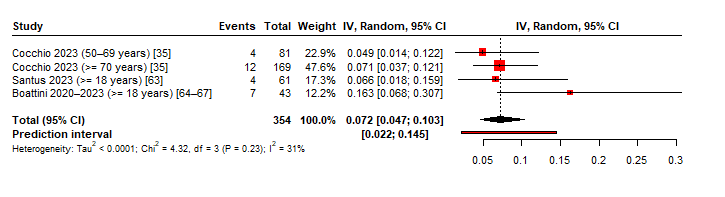
