## Supplementary Table 2 for "Epidemiology and burden of respiratory syncytial virus in Italian adults: A systematic review and meta-analysis"

**S2 Table.** Algorithm for the automatic search, by citation database.

| MEDLINE, Biological Abstracts and Global Health via Ovid | 1 (respiratory syncytial or rsv).mp  2 exp Respiratory Syncytial Viruses/ or exp Respiratory Syncytial Virus, Human/ or exp Respiratory Syncytial Virus Infections/  3 1 or 2  4 exp Adult/ or exp Aging/  5 Men/  6 Women/  7 Retirement/  8 ((old* or age*) adj3 (people* or person* or adult* or women* or men* or citizen* or residen*)).tw.  9 (pension* or retire* or adult* or aged or elderly or senior* or geriatric*).tw.  10 long-term care/ or nursing care/ or palliative care/  11 homes for the aged/ or nursing homes/  12 nursing home*.tw.  13 or/4-12  14 exp Italy/ or (Italy or Italian*).mp or (Italy or Italian*).tw  15 3 and 13 and 14 |
| --- | --- |
| Scopus | TITLE-ABS-KEY ({respiratory syncytial} OR rsv*) AND ALL (adult* OR elder* OR older*) AND TITLE-ABS-KEY (Italy OR Italian*) |
| Web of Science | 1 TS=(respiratory syncytial* or rsv*)  2 TS=(Italy* or Italian*)  3 TS=(adult* or aged or elderly or senior* or geriatric* or retire* or pension* or old* people or old* person* or old* adult* or old* men or old* women or old* citizen* or old* residen* or nursing home*)  4 #1 AND #2 AND #3 |
