## Supplementary Table 3 for "Epidemiology and burden of respiratory syncytial virus in Italian adults: A systematic review and meta-analysis"

**S3 Table.** Excluded studies with reasons.

| **Study** | **Reason for exclusion** |
| --- | --- |
| Rezza 2006 | No separate data on RSV in adults |
| Maggi 2007 | Insufficient data on RSV in adults |
| Montieri 2007 | Children only |
| Debiaggi 2010 | Insufficient data on RSV in adults |
| De Florentiis 2011 | Insufficient data on RSV |
| Cusi 2011 | Children only |
| Boattini 2020 | Analysis of a subset of subjects from a study included (see Boattini et al. [64–66)] |
| Bruyndonckx 2020 | Multi-country study with no separate information for Italy |
| Giardina 2022 | Insufficient data on RSV in adults |
| Duclos 2022 | Multi-country study with no separate information for Italy |
| Amodio 2023 | No separate data for adults and/or RSV |
| Tramuto 2023 | No separate data on RSV in adults |

Rezza G, Valdarchi C, Puzelli S, Ciotti M, Farchi F, Fabiani C, et al. Respiratory viruses and influenza-like illness: a survey in the area of Rome, winter 2004-2005. Euro Surveill. 2006;11(10):251-3.

Maggi F, Andreoli E, Pifferi M, Meschi S, Rocchi J, Bendinelli M. Human bocavirus in Italian patients with respiratory diseases. J Clin Virol. 2007;38(4):321-5. doi: 10.1016/j.jcv.2007.01.008.

Montieri S, Puzelli S, Ciccozzi M, Calzoletti L, Di Martino A, Milia MG, et al. Amino acid changes in the attachment G glycoprotein of human respiratory syncytial viruses (subgroup A) isolated in Italy over several epidemics (1997-2006). J Med Virol. 2007;79(12):1935-42. doi: 10.1002/jmv.21012.

Debiaggi M, Canducci F, Brerra R, Sampaolo M, Marinozzi MC, Parea M, et al. Molecular epidemiology of KI and WU polyomaviruses in infants with acute respiratory disease and in adult hematopoietic stem cell transplant recipients. J Med Virol. 2010;82(1):153-6.

De Florentiis D, Parodi V, Orsi A, Rossi A, Altomonte F, Canepa P, et al. Impact of influenza during the post-pandemic season: epidemiological picture from syndromic and virological surveillance. J Prev Med Hyg. 2011;52(3):134-6.

Boattini M, Almeida A, Christaki E, Cruz L, Antão D, Moreira MI, et al. Influenza and respiratory syncytial virus infections in the oldest-old continent. Eur J Clin Microbiol Infect Dis. 2020;39(11):2085-2090. doi: 10.1007/s10096-020-03959-9.

Cusi MG, Terrosi C, Kleines M, Schildgen O. RSV and HMPV seroprevalence in Tuscany (Italy) and North-Rhine Westfalia (Germany) in the winter season 2009/2010. Influenza Other Respir Viruses. 2011;5(6):380-1. doi: 10.1111/j.1750-2659.2011.00252.x.

Bruyndonckx R, Coenen S, Butler C, Verheij T, Little P, Hens N, et al. Respiratory syncytial virus and influenza virus infection in adult primary care patients: Association of age with prevalence, diagnostic features and illness course. Int J Infect Dis. 2020;95:384-390. doi: 10.1016/j.ijid.2020.04.020.

Giardina FAM, Piralla A, Ferrari G, Zavaglio F, Cassaniti I, Baldanti F. Molecular epidemiology of rhinovirus/enterovirus and their role on cause severe and prolonged infection in hospitalized patients. Microorganisms. 2022;10(4):755. doi: 10.3390/microorganisms10040755.

Duclos M, Hommel B, Allantaz F, Powell M, Posteraro B, Sanguinetti M. Multiplex PCR detection of respiratory tract infections in SARS-CoV-2-negative patients admitted to the emergency department: An international multicenter study during the COVID-19 pandemic. Microbiol Spectr. 2022;10(5):e0236822. doi: 10.1128/spectrum.02368-22.

Amodio E, Vitale F, d'Angela D, Carrieri C, Polistena B, Spandonaro F, et al. Increased risk of hospitalization for pneumonia in Italian adults from 2010 to 2019: Scientific evidence for a call to action. Vaccines (Basel). 2023;11(1):187. doi: 10.3390/vaccines11010187.

Tramuto F, Maida CM, Mazzucco W, Costantino C, Amodio E, Sferlazza G, et al. Molecular epidemiology and genetic diversity of human respiratory syncytial virus in Sicily during pre- and post-COVID-19 surveillance seasons. Pathogens. 2023;12(9):1099. doi: 10.3390/pathogens12091099.
