## Supplementary Figure 4 for "Epidemiology and burden of respiratory syncytial virus in Italian adults: A systematic review and meta-analysis"

**S4 Fig.** Prevalence of RSV subtype B among Italian adults of any age (prevalence of RSV subtype A may be computed as 1 – prevalence of RSV B).

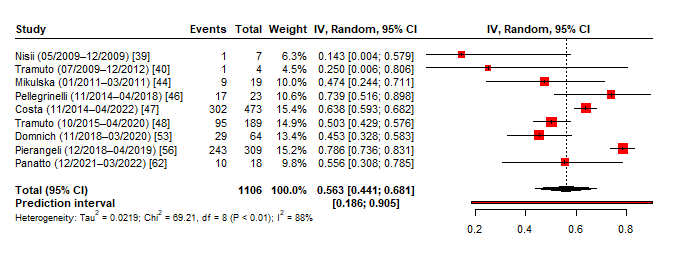
