## Supplementary Table 4 for "Epidemiology and burden of respiratory syncytial virus in Italian adults: A systematic review and meta-analysis"

**S4 Table.** Syndromic definitions used the studies analyzed.

| **Study [Ref]** | **Clinical syndrome** | **Definition** |
| --- | --- | --- |
| Rovida 2005 [30] | ARI | Not otherwise defined. |
| Puzzelli 2009 [32] | ILI | Fever >37.5 °C (axillary temperature) and at least another constitutional symptom (headache, malaise, myalgia, chills, or sweating, retrosternal pain, or asthenia) and one respiratory symptom (cough, sore throat, nasal congestion, or runny nose). |
| Gerna 2009 [34] | ARI | Not otherwise defined. |
| Paba 2014 [37] | ILI | Fever >37.5 °C and at least general symptom (headache, malaise, myalgia, thrill or sweats, retrosternal pain, asthenia) and one respiratory symptom (cough, sore throat, dyspnea). |
| Nisii 2010 [39] | ILI | Abrupt onset of fever (>38°C) with headache or malaise accompanied by either cough, sore throat, or nasal congestion |
| Tramuto 2016 [40] | ILI/ARI with ICU admission | ILI: sudden onset of symptoms and at least one of the four systemic symptoms (fever/feverishness, malaise, headache, myalgia) and at least one of the three respiratory symptoms (cough, sore throat, shortness of breath);  ARI: sudden onset of symptoms and at least one of the four respiratory symptoms (cough, sore throat, shortness of breath, coryza) and a clinician's judgement that the illness is due to an infection. |
| Ansaldi 2012 [42] | ILI | Fever > 38 °C and at least one general symptom (headache, malaise, myalgia, chills or sweats, asthenia) and one respiratory symptom (cough, sore throat, nasal congestion or runny nose). |
| Mikulska 2014 [44] | ILI/ARI | ILI: fever plus at least one respiratory symptom (cough, sore throat, rhinorrhea) and/or systemic symptoms as headache, asthenia, malaise and arthromyalgia, in the absence of other documented causes.  ARI: new onset of symptoms and at least one respiratory symptom (cough, coryza, sore throat, shortness of breath) and a clinician's judgment that the illness is due to an infection. |
| Passi 2019 [45] | URTI | Not otherwise defined. |
| Pellegrinelli 2020 [46] | ILI | Sudden onset of symptoms and at least one of the four systemic symptoms (fever/feverishness, malaise, headache, myalgia) and at least one of the three respiratory symptoms (cough, sore throat, shortness of breath). |
| Tramuto 2021 [48] | ILI/ARI with ICU admission | ILI: sudden onset of symptoms and at least one of the four systemic symptoms (fever/feverishness, malaise, headache, myalgia) and at least one of the three respiratory symptoms (cough, sore throat, shortness of breath);  SARI: hospitalized patients with at least one respiratory (cough, sore throat, or shortness of breath) sign or symptom at admission or within 48 h following admission and at least one systemic (fever or feverishness, malaise, headache, myalgia) sign or symptom or deterioration of general conditions (asthenia, weight loss, anorexia, confusion, dizziness). |
| Leli 2021 [49] | Respiratory infection | Not otherwise defined. |
| De Francesco 2021 [51] | SARI | Hospitalization and the presence of one or more respiratory  symptoms such as shortness of breath, sore throat, cough and fever ≥37.5 °C. |
| Galli 2020 [52] | ILI | Sudden onset of symptoms and at least one of the four systemic symptoms (fever/feverishness, malaise, headache, myalgia) and at least one of the three respiratory symptoms (cough, sore throat, shortness of breath). |
| Domnich 2024 [53] | ILI | Sudden onset of symptoms and at least one of the four systemic symptoms (fever/feverishness, malaise, headache, myalgia) and at least one of the three respiratory symptoms (cough, sore throat, shortness of breath). |
| Sberna 2022 [55] | SARI | Not otherwise defined. |
| Galli 2021 [58] | ILI | Sudden onset of symptoms and at least one of the four systemic symptoms (fever/feverishness, malaise, headache, myalgia) and at least one of the three respiratory symptoms (cough, sore throat, shortness of breath). |
| Milano 2023 [61] | SARI | Hospitalized patients with at least one respiratory (cough, sore throat, shortness of breath) sign or symptom and at least one systemic (fever/feverishness, malaise, headache, myalgia) sign or symptom or deterioration of general conditions (asthenia, weight loss, anorexia, confusion, dizziness) at admission or within 48 h following admission. |
| Santus 2023 [63] | ILI | Any of the following signs or symptoms: cough, sputum, coryza, fever, headache, sore throat, myalgia, asthenia. |

ARI, Acute respiratory infection; ICU, Intensive care unit; ILI, Influenza-like illness; SARI, Severe acute respiratory infection; URTI, Upper respiratory tract infection.
