## Supplementary Table 5 for "Epidemiology and burden of respiratory syncytial virus in Italian adults: A systematic review and meta-analysis"

**S5 Table.** Risk of bias of the studies included according to the Joanna Briggs Institute (JBI) checklist for prevalence/incidence studies.

| **Study** | **Item** | | | | | | | |
| --- | --- | --- | --- | --- | --- | --- | --- | --- |
|  | **1** | **2** | **3** | **4** | **5** | **6** | **7** | **8** |
| Rovida 2005 [30] | Unclear | Yes | No | Yes | Yes | Yes | Yes | Yes |
| Puzzelli 2009 [32] | Yes | Yes | Yes | Yes | Yes | Yes | Yes | Yes |
| Costa 2007 [33] | No | Yes | No | Yes | Yes | Yes | Yes | Yes |
| Gerna 2009 [34] | Yes | Unclear | Yes | Yes | Yes | Yes | Yes | Yes |
| Cocchio 2023 [35] | Yes | Yes | Yes | Yes | Yes | No | Yes | Yes |
| Gambarino 2009 [36] | Yes | Yes | Yes | No | Yes | Yes | Yes | Yes |
| Paba 2014 [37] | Unclear | Unclear | Yes | No | Yes | Unclear | Yes | Yes |
| Pierangeli 2011 [38] | Yes | Yes | Yes | Yes | Yes | Yes | Yes | Yes |
| Tramuto 2016 [40] | Yes | Yes | Yes | Yes | Yes | Yes | Yes | Yes |
| Piralla 2017 [41] | Yes | Yes | Yes | Yes | Yes | Yes | Yes | Yes |
| Ansaldi 2012 [42] | Yes | Yes | Yes | Yes | No | Yes | Yes | Yes |
| Mikulska 2014 [44] | Yes | Yes | Yes | Yes | Yes | Yes | Yes | Yes |
| Pellegrinelli 2020 [46] | Yes | Yes | Yes | Yes | Yes | Yes | Yes | Yes |
| Tramuto 2021 [48] | Yes | Yes | Yes | Yes | Yes | Yes | Yes | Yes |
| Leli 2021 [49] | Unclear | Unclear | Yes | No | Yes | Yes | Yes | Yes |
| Ciotti 2020 [50] | Unclear | Unclear | Yes | Yes | Yes | Yes | Yes | Yes |
| De Francesco 2021 [51] | Yes | Yes | Yes | Yes | Yes | Yes | Yes | Yes |
| Galli 2020 [52] | Unclear | Unclear | Yes | Yes | Yes | Yes | Yes | Yes |
| Domnich 2024 [53] | Yes | Yes | Yes | Yes | Yes | Yes | Yes | Yes |
| Spagnolello 2021 [54] | No | Unclear | No | Yes | Yes | Yes | Yes | Yes |
| Sberna 2022 [55] | Yes | Yes | Yes | No | Yes | Yes | Yes | Yes |
| Pierangeli 2022 [56] | Yes | Yes | Yes | No | Yes | Yes | Yes | Yes |
| Galli 2021 [58] | Yes | Yes | Yes | Yes | Yes | Yes | Yes | Yes |
| Treggiari 2022 [59] | Yes | Unclear | Yes | No | Yes | Yes | Yes | Yes |
| Calderaro 2021 [60] | Yes | Yes | Yes | Yes | Yes | Yes | Yes | Yes |
| Milano 2023 [61] | Yes | Yes | Yes | Yes | Yes | Yes | Yes | Yes |
| Panatto 2023 [62] | Yes | Yes | Yes | Yes | Yes | Yes | Yes | Yes |
| Santus 2023 [63] | Yes | Yes | Yes | Yes | Yes | Yes | Yes | Yes |
| Boattini 2020–2023 [64–66] | Yes | Yes | Yes | Yes | Yes | Yes | Yes | Yes |

**Item legend:**

1. Sample frame appropriate to address the target population?
2. Were study participants recruited in an appropriate way?
3. Was the sample size adequate?
4. Were the study subjects and setting described in detail?
5. Was data analysis conducted with sufficient coverage of the identified sample?
6. Were valid methods used for the identification of the condition?
7. Was the condition measured in a standard, reliable way for all participants?
8. Was there appropriate statistical analysis?
