## Supplementary Table 6 for "Epidemiology and burden of respiratory syncytial virus in Italian adults: A systematic review and meta-analysis"

**S6 Table.** RSV positivity prevalence among Italian adults of any age, by setting.

| **Setting** | **Period** | **Age, years** | **RSV, % (n/N)** | **Ref** |
| --- | --- | --- | --- | --- |
| Outpatient | 11/2004–04/2007 | ≥14 | 1.1 (4/356) | [32] |
|  | 11/2010–04/2011 | ≥60 | 4.4 (2/45) | [42] |
|  | 11/2014–04/2015 | ≥16 | 5.9 (12/203) | [46] |
|  | 11/2015–04/2016 | ≥16 | 8.2 (15/184) | [46] |
|  | 11/2016–04/2017 | ≥16 | 8.2 (15/182) | [46] |
|  | 11/2017–04/2018 | ≥16 | 8.8 (12/137) | [46] |
|  | 11/2018–04/2019 | ≥18 | 8.8 (23/262) | [52] |
|  | 11/2018–04/2019 | ≥18 | 4.1 (30/724) | [53] |
|  | 11/2019–04/2020 | ≥15 | 3.0 (12/401) | [58] |
|  | 11/2019–03/2020 | ≥18 | 6.6 (34/516) | [53] |
|  | 12/2021–03/2022 | ≥18 | 1.6 (19/1213) | [62] |
| Immunocompromised patients | 05/2005–10/2005 | ≥17 | 2.2 (1/46) | [33] |
|  | 11/2006–2005/07 | ≥18 | 8.1 (12/149) | [34] |
|  | 2010–2014 | ≥15 | 3.5 (5/144) | [43] |
|  | 01/2011–03/2011 | ≥18 | 10.9 (21/193) | [44] |
|  | 01/2011–03/2019 | ≥18 | 27.8 (42/151) | [45] |
|  | 11/2019–03/2020 | >18 | 4.4 (8/183) | [57] |
| Inpatient | 10/2001–05/2002 | ≥16 | 5.5 (4/73) | [30] |
|  | 04/2004–05/2005 | ≥18 | 0.2 (1/433) | [31] |
|  | Presumably 2008 | ≥16 | 0.3 (1/324) | [36] |
|  | 07/2009–12/2012 | ≥15 | 2.1 (4/192) | [40] |
|  | 11/2009–04/2016 | ≥18 | 2.9 (11/376) | [41] |
|  | 10/2016–03/2020 | ≥18 | 3.2 (17/539) | [50] |
|  | 01/2017–02/2020 | ≥18 | 7.3 (238/3240) | [51] |
|  | 01/2019–02/2019 | >18 | 4.0 (3/75) | [54] |
|  | 10/2019–12/2019 | ≥18 | 0.8 (1/132) | [55] |
|  | 03/2020–05/2021 | ≥18 | 1.4 (10/734) | [51] |
|  | 10/2020–12/2020 | ≥18 | 0 (0/237) | [55] |
|  | 11/2021–04/2022 | ≥18 | 1.6 (2/129) | [61] |
|  | 10/2021–12/2021 | ≥18 | 4.8 (9/188) | [55] |
|  | 10/2022–03/2023 | ≥18 | 8.5 (61/717) | [63] |
| Mixed | 11/2006–2005/07 | ≥18 | 3.2 (3/95) | [34] |
|  | 02/2009–05/2011 | ≥16 | 16.9 (30/178) | [37] |
|  | 02/2009–03/2010 | ≥18 | 1.7 (4/238) | [38] |
|  | 05/2009–12/2009 | ≥16 | 1.3 (7/544) | [39] |
|  | 11/2014–04/2022 | ≥18 | 4.1 (473/11658) | [47] |
|  | 10/2015–04/2020 | >18 | 4.0 (149/3727) | [48] |
|  | 01/2016–06/2020 | ≥18 | 4.5 (17/375) | [49] |
|  | 12/2019–03/2020 | ≥18 | 2.4 (8/332) | [60] |
|  | 11/2019–03/2020 | ≥60 | 4.1 (3/73) | [59] |
|  | 11/2020–04/2021 | >18 | 0.0 (3/16500) | [59] |
|  | 10/2021–01/2022 | >18 | 2.0 (241/11967) | [59] |
