## Supplementary Table 7 for "Epidemiology and burden of respiratory syncytial virus in Italian adults: A systematic review and meta-analysis"

**S7 Table.** RSV positivity prevalence among Italian working-age adults, by setting.

| **Setting** | **Period** | **Age, years** | **RSV, % (n/N)** | **Ref** |
| --- | --- | --- | --- | --- |
| Outpatient | 11/2004–04/2007 | 14–64 | 1.3 (4/319) | [32] |
|  | 11/2014–04/2015 | 16–65 | 5.4 (9/166) | [46] |
|  | 11/2015–04/2016 | 16–65 | 7.1 (11/154) | [46] |
|  | 11/2016–04/2017 | 16–65 | 6.8 (10/147) | [46] |
|  | 11/2017–04/2018 | 16–65 | 7.9 (9/114) | [46] |
|  | 11/2018–04/2019 | 18–64 | 7.6 (10/131) | [52] |
|  | 11/2018–04/2019 | 18–59 | 3.2 (15/468) | [53] |
|  | 11/2019–04/2020 | 15–64 | 2.4 (8/338) | [58] |
|  | 11/2019–03/2020 | 18–59 | 6.9 (23/334) | [53] |
|  | 12/2021–03/2022 | 18–64 | 1.7 (11/644) | [62] |
| Inpatient | 07/2009–12/2012 | 15–64 | 0 (0/117) | [40] |
|  | 10/2019–12/2019 | 18–64 | 1.3 (1/77) | [55] |
|  | 10/2020–12/2020 | 18–64 | 0 (0/133) | [55] |
|  | 11/2021–04/2022 | 18–64 | 4.3 (2/47) | [61] |
|  | 10/2021–12/2021 | 18–64 | 3.1 (3/97) | [55] |
|  | 10/2022–03/2023 | 18–69 | 6.6 (18/273) | [63] |
| Mixed | 11/2014–04/2022 | 18–59 | 3.2 (136/4239) | [47] |
|  | 10/2015–04/2020 | 19–64 | 3.6 (95/2674) | [48] |
|  | 01/2016–06/2020 | 18–64 | 2.0 (4/200) | [49] |
|  | 11/2020–04/2021 | 19–59 | 0.0 (3/10197) | [59] |
|  | 10/2021–01/2022 | 19–59 | 2.3 (163/7154) | [59] |
