## Supplementary Table 8 for "Epidemiology and burden of respiratory syncytial virus in Italian adults: A systematic review and meta-analysis"

**S8 Table.** RSV positivity prevalence among Italian older adults, by setting.

| **Setting** | **Period** | **Age, years** | **RSV, % (n/N)** | **Ref** |
| --- | --- | --- | --- | --- |
| Outpatient | 11/2004–04/2007 | ≥65 | 0 (0/37) | [32] |
|  | 11/2010–04/2011 | ≥60 | 4.4 (2/45) | [42] |
|  | 11/2014–04/2015 | >65 | 8.1 (3/37) | [46] |
|  | 11/2015–04/2016 | >65 | 13.3 (4/30) | [46] |
|  | 11/2016–04/2017 | >65 | 14.3 (5/35) | [46] |
|  | 11/2017–04/2018 | >65 | 13.0 (3/23) | [46] |
|  | 11/2018–04/2019 | ≥60 | 5.9 (15/256) | [53] |
|  | 11/2019–04/2020 | ≥65 | 6.3 (4/63) | [58] |
|  | 11/2019–03/2020 | ≥60 | 6.0 (11/182) | [53] |
|  | 12/2021–03/2022 | ≥65 | 1.4 (8/569) | [62] |
| Inpatient | 07/2009–12/2012 | ≥65 | 5.3 (4/75) | [40] |
|  | 10/2019–12/2019 | ≥65 | 0 (0/55) | [55] |
|  | 10/2020–12/2020 | ≥65 | 0 (0/104) | [55] |
|  | 11/2021–04/2022 | ≥65 | 0 (0/82) | [61] |
|  | 10/2021–12/2021 | ≥65 | 6.6 (6/91) | [55] |
|  | 10/2022–03/2023 | ≥70 | 9.7 (43/444) | [63] |
| Mixed | 11/2014–04/2022 | ≥60 | 4.5 (337/7419) | [47] |
|  | 10/2015–04/2020 | ≥65 | 5.1 (54/1053) | [48] |
|  | 01/2016–06/2020 | ≥65 | 7.4 (13/175) | [49] |
|  | 11/2019–03/2020 | ≥60 | 4.1 (3/73) | [59] |
|  | 11/2020–04/2021 | ≥60 | 0 (0/6303) | [59] |
|  | 10/2021–01/2022 | ≥60 | 1.6 (78/4813) | [59] |
