## Supplementary Table 9 for "Epidemiology and burden of respiratory syncytial virus in Italian adults: A systematic review and meta-analysis"

**S9 Table.** Meta-regression analysis to investigate sources of heterogeneity in RSV positivity prevalence among Italian adults of any age.

| **Variable** | **Level** | **Coefficient** | **Standard error** | **P-value** |
| --- | --- | --- | --- | --- |
| Year of publication | 1-year increase | 0.003 | 0.003 | 0.35 |
| Public funding | No | Ref | Ref | Ref |
|  | Yes | 0.008 | 0.037 | 0.82 |
| Geographic area | North | Ref | Ref | Ref |
|  | Center | -0.072 | 0.035 | 0.041 |
|  | South | -0.066 | 0.069 | 0.34 |
| Setting | Inpatient | Ref | Ref | Ref |
|  | Outpatient | 0.052 | 0.037 | 0.16 |
|  | Mixed | 0.021 | 0.038 | 0.59 |
|  | Immunocompromised patients | 0.169 | 0.048 | < 0.001 |
| Multi-season study | No | Ref | Ref | Ref |
|  | Yes | 0.041 | 0.035 | 0.24 |
| Study period | Before COVID-19 pandemic | Ref | Ref | Ref |
|  | During COVID-19 pandemic | -0.035 | 0.041 | 0.40 |
| Out-of-season samples | No | Ref | Ref | Ref |
|  | Yes | 0.007 | 0.033 | 0.84 |
| Specimen type | Lower respiratory tract | Ref | Ref | Ref |
|  | Upper respiratory tract | 0.067 | 0.058 | 0.25 |
|  | Both | 0.078 | 0.062 | 0.21 |
| Proportion of older adults | <50% | Ref | Ref | Ref |
|  | >50% | 0.068 | 0.037 | 0.067 |
| Sample size | >300 | Ref | Ref | Ref |
|  | <300 | 0.087 | 0.029 | 0.003 |
| Risk of bias | Lower | Ref | Ref | Ref |
|  | Higher | 0.003 | 0.03 | 0.92 |
