## Supplementary Table 10 for "Epidemiology and burden of respiratory syncytial virus in Italian adults: A systematic review and meta-analysis"

**S10 Table.** RSV positivity prevalence among Italian adults, by subtype.

| **Period** | **RSV A, % (n/N)** | **RSV B, % (n/N)** | **Ref** |
| --- | --- | --- | --- |
| 05/2009–12/2009 | 85.7 (6/7) | 14.3 (1/7) | [39] |
| 07/2009–12/2012 | 75.0 (3/4) | 25.0 (1/4) | [40] |
| 01/2011–03/2011 | 52.6 (10/19) | 47.4 (9/19) | [44] |
| 11/2014–04/2018 | 26.1 (6/23) | 73.9 (17/23) | [46] |
| 11/2014–04/2022 | 36.2 (171/473) | 63.8 (302/473) | [47] |
| 10/2015–04/2020 | 28.6 (54/189) | 71.4 (95/189) | [48] |
| 11/2018–03/2020 | 54.7 (35/64) | 45.3 (29/64) | [53] |
| 12/2018–04/2019 | 21.4 (66/309) | 78.6 (243/309) | [56] |
| 12/2021–03/2022 | 44.4 (8/18) | 55.6 (10/18) | [62] |
