## Supplementary Table 11 for "Epidemiology and burden of respiratory syncytial virus in Italian adults: A systematic review and meta-analysis"

**S11 Table.** Frequency of viral co-detections among RSV-positive Italian adults of any age.

| **N RSV positive samples** | **Period** | **Co-detection, % (n/N)** | **Co-detected pathogens (n)** | **Ref** |
| --- | --- | --- | --- | --- |
| ≥30 | 02/2009–05/2011 | 3.3 (1/30) | AdV (1) | [37] |
|  | 11/2014–04/2022 | 18.6 (88/473) | FluA (27), CoV (20), HRV (20), MPV (7), PIV-3 (5), FluB (2), AdV (2), PIV-1 (1), PIV-2 (1), PIV-3 + BoV (1), AdV + HRV (1), CoV + HRV (1) | [47] |
|  | 01/2017–02/2020 | 2.9 (7/238) | CoV (2), HRV (1), FluA (1), FluB (1), PIV (1), AdV (1) | [51] |
|  | 11/2018–03/2020 | 14.1 (9/64) | FluA (2), FluB (2), HRV (2), CoV (1), PIV (1), AdV (1) | [53] |
|  | 10/2022–03/2023 | 6.6 (4/61) | MPV (2), FluB (2) | [63] |
| <30 | 10/2001–05/2002 | 100 (4/4) | PIV (2), FluA (1), FluB (1) | [30] |
|  | 04/2004–05/2005 | 100 (1/1) | HRV (1) | [31] |
|  | Presumably 2008 | 0 (0/1) | – | [36] |
|  | 07/2009–12/2012 | 25.0 (1/4) | CoV-229E (1) | [40] |
|  | 01/2019–02/2020 | 33.3 (1/3) | BoV (1) | [54] |
|  | 12/2019–03/2020 | 25.0 (2/8) | HRV (1), CoV (1) | [60] |
|  | 03/2020–05/2021 | 0 (0/10) | – | [51] |
|  | 11/2021–04/2022 | 50.0 (1/2) | SARS-CoV-2 (1) | [61] |
|  | 12/2021–03/2022 | 52.6 (10/19) | SARS-CoV-2 (6), HEV (1), HRV (1), SARS-CoV-2 + HRV (1), CoV-229E (1) | [62] |

AdV, Adenovirus; BoV, Bocavirus; CoV, Seasonal coronavirus; FluA, Influenza type A virus; FluB, Influenza type A virus; HRV, Human rhinovirus; MPV, Metapneumovirus; PIV, Parainfluenza virus.
